# Molecular epidemiology of *Escherichia coli* and *Klebsiella* species bloodstream infections in Oxfordshire (UK) 2008-2018

**DOI:** 10.1101/2021.01.05.20232553

**Authors:** Samuel Lipworth, Karina-Doris Vihta, Kevin Chau, Leanne Barker, Sophie George, James Kavanagh, Timothy Davies, Alison Vaughan, Monique Andersson, Katie Jeffery, Sarah Oakley, Marcus Morgan, Susan Hopkins, Timothy E. A. Peto, Derrick W. Crook, Ann Sarah Walker, Nicole Stoesser

**Affiliations:** Nuffield Department of Medicine, University of Oxford, Oxford, United Kingdom; Oxford University Hospitals NHS Foundation Trust, Oxford, United Kingdom; NIHR Health Protection Research Unit in Healthcare Associated Infections and Antimicrobial Resistance at University of Oxford in partnership with Public Health England, Oxford, United Kingdom; National Infection Service, Public Health England, Colindale, London; NIHR Biomedical Research Centre, Oxford, United Kingdom

**Keywords:** Gram-negative bloodstream infections, Bacteraemia, Whole genome sequencing, *Klebsiella pneumoniae*, virulence, antimicrobial resistance

## Abstract

The incidence of Gram-negative bloodstream infections (BSIs), predominantly caused by *Escherichia coli* and *Klebsiella* species, continues to increase; however the causes of this are unclear and effective interventions are therefore hard to design. In this study we sequenced 3468 sequential, unselected isolates over a decade in Oxfordshire, UK. We demonstrate that the observed increases in *E. coli* incidence were not driven by clonal expansion; instead, four major sequence types (STs) continue to dominate a stable population structure, with no evidence of adaptation to hospital/community settings. Conversely in *Klebsiella* spp. most infections are caused by sporadic STs with the exception of a local drug-resistant outbreak strain (ST490). Virulence elements are highly structured by ST in *E. coli* but not *Klebsiella* spp. where they occur in a diverse spectrum of STs and equally across healthcare and community settings. Most clinically hypervirulent (i.e. community-onset) Klebsiella BSIs have no known acquired virulence loci. Finally we demonstrate a diverse but largely genus-restricted mobilome with close associations between antimicrobial resistance (AMR) genes and insertion sequences but not typically specific plasmid replicon types; consistent with the dissemination of AMR genes being highly contingent on smaller mobile genetic elements (MGEs). Our large genomic study highlights distinct differences in the molecular epidemiology of *E. coli* and *Klebsiella* BSIs, and suggests that no single specific pathogen genetic factors are likely contributing to the increasing incidence of BSI overall, that association with AMR genes in *E. coli* is a contributor to the increasing number of *E. coli* BSIs, and that more attention should be given to AMR gene associations with non-plasmid MGEs to try and understand horizontal gene transfer networks.

## Background

Gram-negative bloodstream infections (GNBSI), predominantly caused by *Escherichia coli* and *Klebsiella* spp., are a significant and increasing threat to public health. They are now the leading cause of bloodstream infection (BSI) in the UK with a substantial associated burden of morbidity and mortality^1,2^. Despite becoming a significant public health concern and policy focus, their incidence continues to increase and it is unclear how ambitious targets to reduce this can be achieved^3^.

There is a significant association between GNBSIs and antimicrobial resistance (AMR), particularly in certain globally successful sequence types (STs), such as ST131^4,5^. Enterobacteriaceae have relatively open pan-genomes and are able to rapidly adapt to changing selection pressures (including antibiotic usage)^6,7^. Multidrug resistance (MDR)-associated STs have been linked with prolonged hospital stay and adverse outcomes^8^. Whilst infections caused by relatively susceptible isolates still represent the majority of cases, the potential for rapid proliferation of AMR-associated clones and the dissemination of AMR genes on mobile genetic elements between lineages and species is a major concern^9^. Recent molecular epidemiology studies in the UK have replicated global findings that most *E. coli* BSIs are caused by the clonal lineages ST131, 95, 73 (all phylogroup B2) and ST 69 (phylogroup D)^7,10^. One study has shown that after the emergence and clonal expansion of ST69 and ST131 in the early 2000s, the population structure reached an equilibrium, with STs 131, 95, 73 and 69 predominating^11^. However, this study only sequenced isolates cultured prior to 2012, which may represent a critical inflection point in the incidence rate,^1^ which continues to increase year on year. It remains unclear whether this is caused by homogenous expansion of *E. coli* lineages or clonal expansion of an existing or emergent ST.

For *Klebsiella* spp., it has been hypothesised that isolates broadly cause two categories of BSI^12,13^, namely: (i) multidrug resistant healthcare-associated (HA) infections caused by low virulence strains with AMR gene-associated plasmids, and (ii) clinically hypervirulent community-associated (CA) infections caused by strains carrying virulence gene plasmids. The convergence of these two phenotypes might pose a significant risk to human health^14^. Whilst there are multiple studies describing in detail the epidemiology of globally distributed clones associated with AMR and hyper-virulence^5,15^, such isolates represent a minority of BSI in the UK and most of Europe^16^. A recent study in Cambridgeshire (UK) of 162 *K. pneumoniae* isolates, enriched to over-represent MDR isolates, demonstrated a predominance of globally important clones with apparent cycling of relative incidence over a two-year period^17^. However, MDR infections represent approximately 18% of *K. pneumoniae* BSIs in England and the molecular epidemiology of most invasive disease caused by *Klebsiella* spp. has not been systematically studied^2^.

In order to investigate underlying potential pathogen genetic factors driving the increasing incidence of these infections, we analysed all Klebsiella *spp*. and *E. coli* BSIs over a decade (2008-2018) in Oxfordshire, UK, linking electronic health records and mandatory reporting data with sequencing data for all isolates, to give a detailed picture of the regional genomic epidemiology of *E. coli* and *Klebsiella spp*. BSIs. We used other publicly available sequencing datasets to contextualise our findings.

## Methods

### Study setting, laboratory procedures and DNA extraction

All isolates causing BSI between 15/09/2008-01/12/2018 (de-duplicated to one BSI/90-day period within-patient) were processed by the clinical microbiology laboratory at the John Radcliffe Hospital, Oxford, UK, using standard operating procedures, with sub-cultured stocks stored at -80°C in 10% glycerol nutrient broth. The microbiology laboratory serves all healthcare facilities in Oxfordshire, UK, with a catchment population of ∼805,000. For sequencing, isolate stocks were grown overnight on Columbia blood agar at 37°C and DNA was extracted using the QuickGene DNA extraction kit (Autogen, MA, USA) as per the manufacturer’s instructions with the addition of a mechanical lysis step (FastPrep, MP Biomedicals, CA, USA; 6m/s for 40 secs). Short-read sequencing was performed using the Illumina HiSeq 2500/3000/4000/MiSeq instruments as previously described^18^. Additional, publicly available short-read sequencing data was obtained to facilitate phylogenetic comparison with previous studies^7,11,19,20^ (BioProject accessions: PRJNA480723, PRJEB4681, PRJEB35000, PRJEB12513).

### Epidemiological linkage

Isolate data was linked to laboratory and electronic health record data via the Infections in Oxfordshire Research Database (IORD). IORD has generic Research Ethics Committee, Health Research Authority and Confidentiality Advisory Group approvals (19/SC/0403, 19/CAG/0144) as a de-identified electronic research database. Data on suspected infectious focus and patient provenance was acquired via linked local infection control records which had been submitted to Public Health England as part of the mandatory surveillance programme; such data was available for 400 *Klebsiella* spp. and 2773 *E. coli* isolates. Healthcare-associated (HA) BSI were defined as occurring >48 hrs post-hospital admission or ≤30 days since hospital discharge; other cases were defined as community-associated (CA)^21^.

### Genomic analysis

All programs were run using default settings unless indicated. Raw reads were assembled using Shovill (v1.0.4) with assemblies <4Mb and >7Mb excluded from further analysis, because these were thought to represent possible assembly errors given the typical genome size for these species (4-6.5Mb)^22^. De novo assemblies were annotated with Prokka (v1.14.6)^23^; AMR genes, virulence factors and plasmid replicons were identified using the Resfinder, VFDB, ISFinder and PlasmidFinder databases with Abricate (v0.9.8) (--min-id 95 --min-cov 95)^24^, and Kleborate for *Klebsiella* spp.^12,25–27^. Multi-locus sequence types (MLST) were determined *in silico* using the MLST tool (v2.17.6) for *E. coli* and Kleborate for *Klebsiella* spp.^28,29^. Phylogroups were determined using the ClermonTyping tool^30^. To fully utilise the resolution provided by whole genome sequencing we additionally used fastBAPS^31^ to partition the population using the core gene alignment produced by Panaroo^32^.

Mapping to MLST-specific reference genomes was performed using Snippy (v4.6.0)^33^; genomes were acquired from NCBI (appendix). Gubbins (v2.3.4) was used to build recombination-corrected phylogenies^34^. Time-scaled phylogenies were created using the BactDating library in R (version 3.6.0) under a relaxed gamma model with a minimum of 100,000 Markov chain Monte Carlo iterations^35^. Significance of temporal signal was assessed with root-to-tip plots and 10,000 random permutations of tip dates. Effective sample size was assessed using the Coda package^36^. Evolutionary distinctiveness (ED) was calculated using the Picante package in R^37^; low ED scores indicate closely related genomes. All bioinformatics was performed using the Oxford University Biomedical Research Computing Facility.

Contigs were classified as being of likely chromosomal or plasmid origin using MLplasmids with a 0.7 probability cut-off^38^. All plasmid/chromosomal contigs for a given isolate were binned into separate multi-fasta files and the distances between these calculated using Dashing^39^. The pairwise distance matrix was filtered on a distance of 0.71 (the median plasmidome similarity of bla_CTX-M-15_ carrying *K. pneumoniae* ST490 isolates) and then clustered into “plasmidome groups” using the LinkComm package in R^40^. The ecology of categorical groups was compared with a permutational multivariate analysis of variance (permanova) test in the Vegan package in R^41^.

A pangenome wide association study (PGWAS) was conducted using PySeer^42^ with a linear mixed model utilising relatedness distances inferred from a core genome phylogeny to correct for population structure. A discriminant analysis of principal components (DAPC) was performed using the Adegenet library in R^43^. Gene co-occurrence was examined using graphs constructed using the iGraph library in R^44^. For each species, edge lists with all genes/plasmids/insertion sequences with a Pearson correlation coefficient spp. >0.5 were created and communities detected using single linkage.

### Statistical analysis

To describe molecular epidemiological trends, STs were arbitrarily categorised as rare (≤10 isolates over the study period, or untypeable STs), intermediate (11-100 isolates), and for *E. coli*, submajor (101-300 isolates) and major (>=300 isolates). For Klebsiella spp. we considered the most prevalent ST (490) in isolation. Stacked negative binomial regression models with clustered standard errors were used to compare rates of change over time (STATA v16^45^)^46^. Incomplete years (2008 and 2018) are excluded from this part of the analysis. All other statistical tests were performed in R v3.6 using the Stats package unless indicated^47^. Charlson score was calculated in the Comorbidity^48^ R package using all ICD10 codes associated with episodes in the year prior to specimen collection dates. To test for geographical/healthcare setting structure in MLST groups we conducted a permutation test similar to that previously described^49^. In brief, the ratio of median distances between isolates from the same centre and different centres was calculated. This observed ratio was compared to a permuted distribution created by 1000 tip label randomisations. The number of permuted values at least as extreme as the observed value divided by the number of permutations carried out was calculated to give a one-sided test of statistical significance.

## Results

### Escherichia coli bloodstream infections are stably dominated by major lineages across community and healthcare-associated settings, but with diverse sporadic lineages accounting for almost half of cases, and evidence of sub-lineage replacement in major STs

From September 2008-December 2018, 3461 *E. coli* isolates from 3196 patients were sequenced. Major STs, namely ST73 (n=574), ST131 (457), ST95 (320) and ST69 (314) comprised 48% of all isolates; sub-major STs, ST12 (144) and ST127 (113), 7% of all isolates; intermediate STs (32 STs) 24% of all isolates; and rare STs (304 STs) 21% of all isolates (Fig.1/Fig.S1). The incidence rate ratio per year (IRRy) of all these categories increased over time (major IRRy=1.13 (95% CI:1.10-1.17), sub-major IRRy=1.16 (1.10-1.22), intermediate IRRy=1.15 (1.12-1.18), rare IRRy=1.10 (1.06-1.14) (Fig.1A). There was no evidence that the proportion of BSIs caused by major or sub-major STs changed over the study period (Fig.1B). There was evidence that the incidence of BSIs caused by intermediate STs increased slightly faster versus all other isolates and by rare isolates slightly slower (p_heterogeneity_<0.05). After sub-stratifying the major STs 69, 73, 95 and 131 using BAPS, there was some evidence of sublineage replacement (Fig.S2/Table.S1).

**Figure 1.**
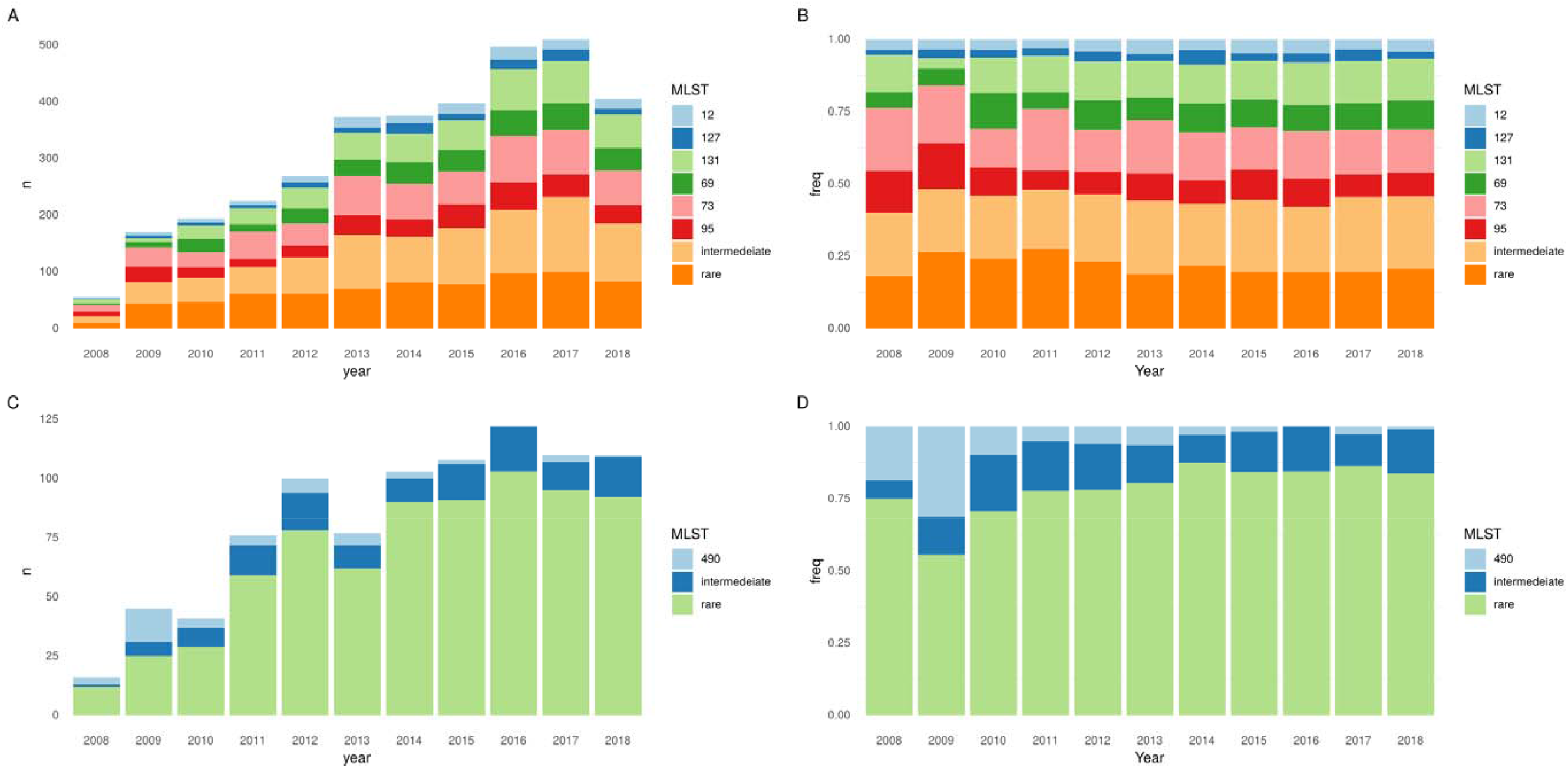
Population dynamics of *E. coli* (A, B) and *Klebsiella* spp. (C, D) STs over time. A) Absolute number and B) proportions of *E. coli* BSIs caused by four major STs (STs 131/95/73/69), sub-major STs (STs 127/12), intermediate STs and rare STs. C) Absolute number and D) proportions of *Klebsiella* s BSIs caused by ST490, intermediate, and rare STs. N.B years 2008 and 2018 are incomplete (see methods).

The majority of *E. coli* BSIs were CA-BSIs (2104/3461, 61%), with no evidence of variation in the proportion caused by major STs between CA-BSI and HA-BSI (990/2104, 47%, vs 661/1357, 49%, p=0.4). Relatively few isolates came from patients resident in a nursing/care home was relatively low (169 (7%) of 2301 with data available). The four major *E. coli* STs accounted for a significantly greater proportion of these cases (97/169, 57% vs 1009/2132, 47% p=0.01); however, there was no evidence of large-scale nursing/care home-associated BSI outbreaks (Fig.S3). E. coli CA-BSIs were slightly older (median age 76 (IQR 63-85) vs 70 (55-80), p<0.001) and less comorbid (median Charlson score 1 (IQR 0-2) vs 1 (IQR 1-2), p=0.003) than HA-BSIs.

### Klebsiella spp. bloodstream infections are caused by a diverse representation of sub-species with significant intra-species diversity, with the exception of K. pneumoniae ST490, causing a transient, clonal, local outbreak

Amongst the 886 successfully sequenced *Klebsiella* spp. isolates, a large number of sub-species were observed, including *K. pneumoniae* (n=528 [60%]), *K. variicola* (n=112 [13%]), *K. michiganensis* (n=90 [10%]), *K. oxytoca* (n=59 [7%]), *K. aerogenes* (n=38 [4%]), *K. grimontii* (n=30 [3%]), *K. quasipneumoniae* (n=28 [3%]) and *K. africana* (n=1 [0.1%]). In stark contrast to *E. coli*, 738/886 (83%) of *Klebsiella* spp. isolates belonged to rare STs (Fig.1/Fig.S4). The multidrug resistant *K. pneumoniae* ST490 was the most prevalent *Klebsiella* ST and the only one with >40 isolates; its incidence decreased over the study period (IRRy 0.78, 95%CI:0.68-0.89). This ST is rarely seen in other studies, consistent with a transient but relatively large, local outbreak. Notably 16/45 (36%) of isolates from this ST were community-onset.

The majority of *Klebsiella* spp. BSIs were HA-BSI (510/882 [missing data for 4 isolates], 59%). As with *E. coli* few *K. pneumoniae* cases were in patients resident in a nursing/care home (10/151, 7%) though this data was not available for most isolates. There was no difference in the proportion of intermediate STs amongst HA-BSI vs CA-BSI (90/510 18% vs 58/372 16%, p=0.5). *Klebsiella* spp. CA-BSI cases were older (median age 76 years (IQR 65-85) vs 66 years (49-75) for HA-BSI; p<0.001) but also less comorbid than HA-BSI cases (median Charlson score 1 (IQR 0-2) vs 2 (1-3); p<0.001). A large number (265/341, missing data for 31) of CA-BSIs occurred in relatively healthy individuals (Charlson score ≤2 i.e. predicted 10 year survival ∼90%). In contrast to previous reports suggesting *K. quasipneumoniae*/*K. variicola* may be less virulent than *K. pneumoniae*^7^, in our study there was no evidence of differences in crude 30-day mortality between the *Klebsiella* subspecies with >= 20 isolates (111/524 (21%) *K. pneumoniae*, 28/112 (25%) *K. variicola*, 23/89 (26%) *K. michiganensis*, 9/57 (16%) *K. oxytoca*, 10/36 (28%) *K. aerogenes*, 7/28 (25%) *K. quasipneumoniae*, and 7/29 (24%) *K. grimontii;* exact p=0.5, missing data for 9 isolates), nor the proportion of Ca-BSI (230/527 (44%), 48/112 (43%), 34/89 (38%), 27/58 (47%), 11/28 (39%), 9/27 (25%) and 12/30 (40%) respectively; exact p=0.4, missing data for 4 isolates).

### No evidence of intra- or inter-regional clustering of major Escherichia coli lineages causing BSI in either community- or healthcare-associated settings

For the four largest *E. coli* STs, we compared the phylogeny of BSI isolates in this study to those in previous studies over a similar timeframe (see Methods). There was no obvious clustering of HA or CA isolates (Fig.2), supported by permutation testing, confirming that for the major STs these were indeed randomly distributed across the phylogeny by geography and healthcare/community setting (Table S2). Additionally, for all four of the major *E. coli* STs, there was no difference in the ED scores between HA and CA cases, and therefore no evidence of healthcare-associated adaptation and clonal expansion (Table S3). Our PGWAS did not identify any significant hits separating CA and HA isolates (Fig.S5), including those that had been previously identified in other studies^47^.

**Figure 2:**
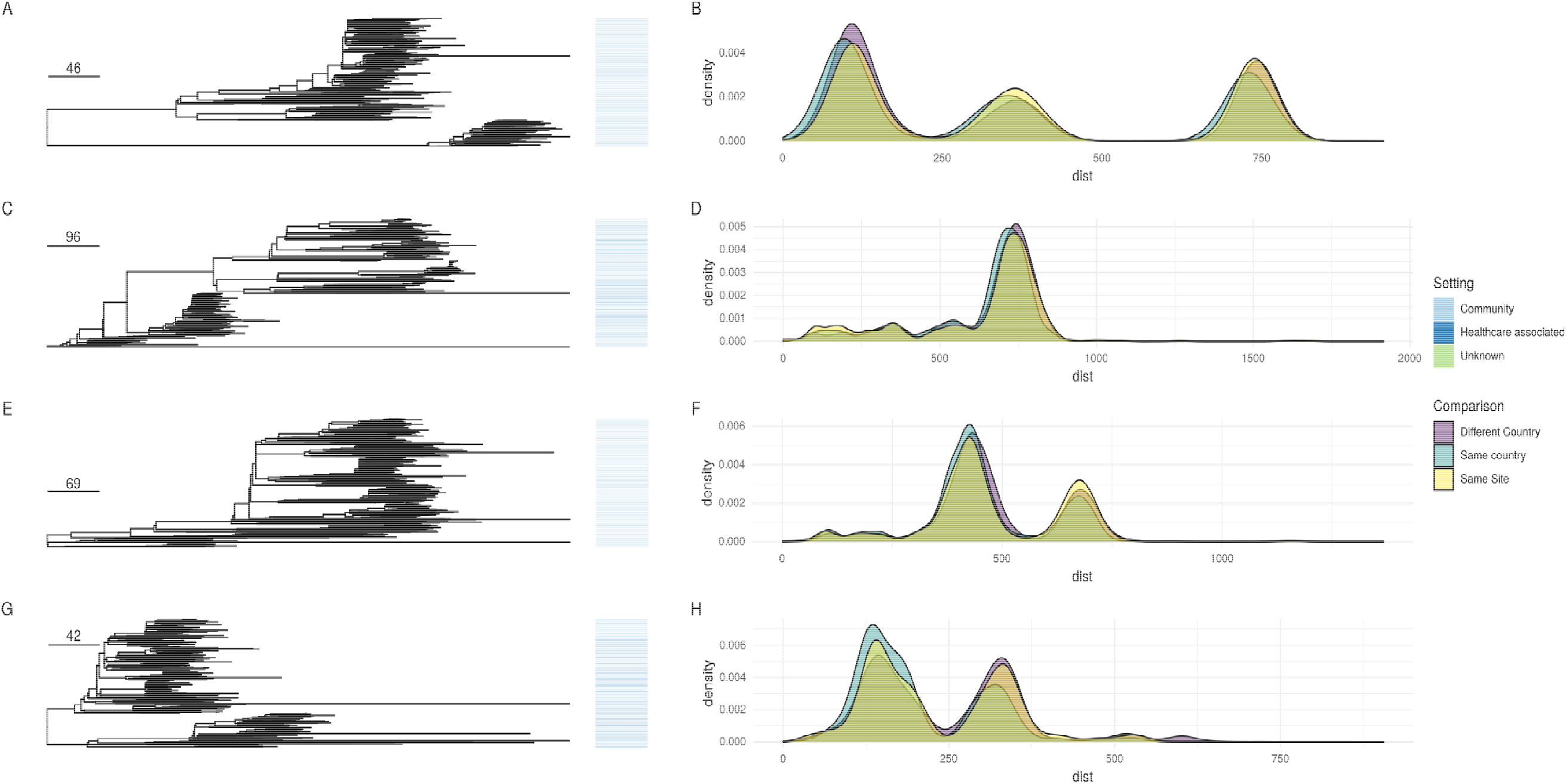
Left panel: Core genome phylogenies (corrected for recombination) for ST131/95/73/69 (top to bottom). Origin of isolates is denoted by the colour in the bar the right of the tree. Scale bar shows SNPs. Right panel shows the distribution of cophenetic distances for the trees shown for isolates from the same site, same countr different countries.

### Urinary and hepatobiliary sources of E. coli and Klebsiella spp. BSI predominate, with major E. coli STs over-represented in urinary-associated E. coli BSIs

The urinary tract was the most common physician-identified source of infection in both species. In *E. coli*, these were strongly associated with the presence of the pap group of proteins (Table S4). The hepatobiliary was the second most common source followed by other gastrointestinal infections for *E. coli* and a respiratory source for *Klebsiella* spp. (Fig.S6a). For *E. coli* BSIs, there was some evidence that incidence of BSIs not attributed to the urinary tract increased faster than those thought to be of urinary origin (IRRy 1.45, 95%CI 0.98-2.17 vs IRRy 1.36, 95%CI 0.88-2.08 respectively p_heterogeneity_=0.005, Fig.S6b). The proportion of BSIs caused by the five major *E. coli* STs was greater for BSIs with urinary compared to other sources (586/1071 [55%], vs 520/1229 [42%], p<0.001); this was not the case for *Klebsiella* spp. where the STs causing BSI attributable to all sources were diverse.

### Multi-drug resistant E. coli BSIs are increasing, likely driven by the association of AMR genes within lineages, and ceftriaxone resistance is driven by the expansion of resistant ST131 and ST73 sub-clades

For E. coli BSIs, considering total effects of resistance to individual antibiotics, regardless of mechanism, there was evidence of increasing incidence of BSI carrying AMR genes conferring resistance to amoxicillin IRRy=1.17 (95%CI 1.14-1.20), co-amoxiclav IRRy=1.15 (1.08-1.24), ceftriaxone IRRy=1.26 (1.17-1.36), gentamicin IRRy=1.24 (1.15-1.33), trimethoprim IRRy=1.17 (1.14-1.20) and ciprofloxacin (IRRy=1.23 (1.17-1.29) Fig.3); there was only a single isolate carrying a gene encoding for carbapenem resistance (ST10, bla_OXA-48_) in 2014. For ceftriaxone (p_heterogeneity_=0.003), gentamicin (p_heterogeneity_=0.03), amoxicillin (p_heterogeneity_=0.003), ciprofloxacin (p_heterogeneity_<0.001), there was evidence that the incidence of isolates with AMR genes encoding resistance to these antimicrobials was increasing faster than those without. Genes conferring resistance to ceftriaxone in E. coli were over-represented in ST131 (180/346) and ST73 (56/346), with significantly lower ED scores for isolates carrying ceftriaxone-resistance genes in these STs (accounting for 236/346 (68%) of these genes, Figs.S1), consistent with clonal expansion of resistant sub-clades within these lineages. Findings were similar restricting to CA-BSI. ST127 was the only major/sub-major ST in which no genes conferring resistance to ceftriaxone were detected.

**Figure 3:**
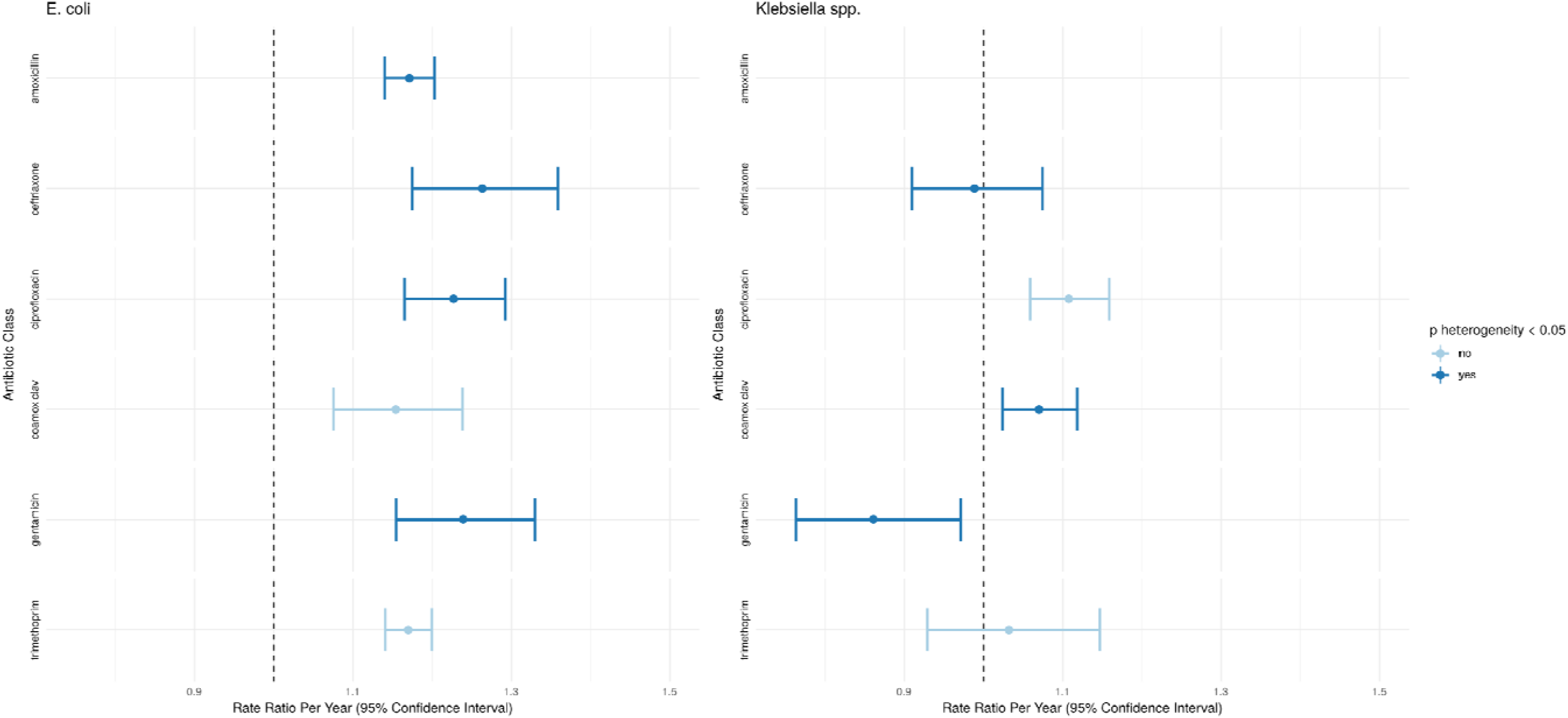
Changes in the incidence of clinically relevant groups of antimicrobial resistance genes considering total effects of resistance to individual antibiotics, regardless of mechanism, over the study period. Gene groupings are as defined by ResFinder. Amoxicillin is excluded for *Klebsiella* spp. which are generally considered intrinsically resistant to this drug. Bars show univariate point estimates and 95% confidence intervals for the incidence rate ratio (per year) of presence of genes belonging to the classes shown. The point estimate and error bar is shown in a darker blue if a Wald test between the incidence rate ratios for the presence and absence of a given set of genes was significant at a 0.05 threshold (i.e. there was evidence that the rate of increase of isolates with a group of genes was different to that for isolates without.)

### Drug-susceptible Klebsiella spp. BSIs are increasing faster than drug-resistant strains, with ceftriaxone-resistance largely healthcare-associated

For *Klebsiella* spp., there was only evidence of increasing incidence of isolates carrying genes/mutations conferring resistance to co-amoxiclav IRRy=1.07 (95%CI:1.02-1.12) and ciprofloxacin IRRy=1.11 (1.06-1.16) (Fig.3). In contrast, the incidence of genes conferring resistance to ceftriaxone IRRy 0.99 (0.92-1.09) and trimethoprim IRRy=1.03 (0.93-1.15) were stable while gentamicin IRRy=0.86 (0.76-0.97) decreased. For ceftriaxone, co-amoxiclav and gentamicin, the incidence trends of isolates not carrying genes conferring resistance to these antibiotics increased faster than those carrying resistance (p_heterogeneity_<=0.01).

A major contributing factor to this finding was the overall decline of the MDR ST490 lineage and the emergence of a more susceptible ST490 sub-lineage between 2005-2010 which had lost the *aac(3)-IIa, aac(6’)-Ib-cr, bla*_OXA-1_ and *tet(a)* genes (Fig.S7). Genes encoding for resistance to ceftriaxone were significantly more common in intermediate STs (STs occurring >10 times in the dataset) (54/148, 36% vs 73/738, 10%, p <0.001) and in HA-BSIs (85/510, 17% vs 42/372, 11% p=0.03). Similarly MDR isolates (resistant to >=3 antibiotic classes) were more common in intermediate vs rare STs (89/366, 24% vs 80/520, 15%, p=0.001) and HA-BSI vs CA-BSI (109/510, 21% vs 59/372, 16%, p=0.049).

### Virulence factors are strongly structured by ST amongst E. coli but not Klebisella spp. BSI isolates with no evidence in Klebsiella of a difference in virulence gene carriage between community vs hospital acquired BSIs

The distribution of virulence factors strongly reflected the underlying clonal population structure in *E. coli* isolates, with segregation by ST, confirmed with a discriminant analysis of principal components (DAPC) (Fig.4). Most notably phylogroup B2 isolates were separated from the rest of the population by the presence of *chuA, chuX* (involved in haem utilisation) and *espL1/espX4* (elements of the type 3 secretion system). Given this and the broad equilibrium in the E. coli BSI population structure demonstrated above, it is unlikely that increased carriage of certain virulence factors explains the increasing incidence of these infections. In keeping with this, there was a near-perfect correlation between frequency of carriage of any given virulence gene across all isolates in 2009 vs 2018 (Pearson correlation=0.99; p<0.001).

**Figure 4:**
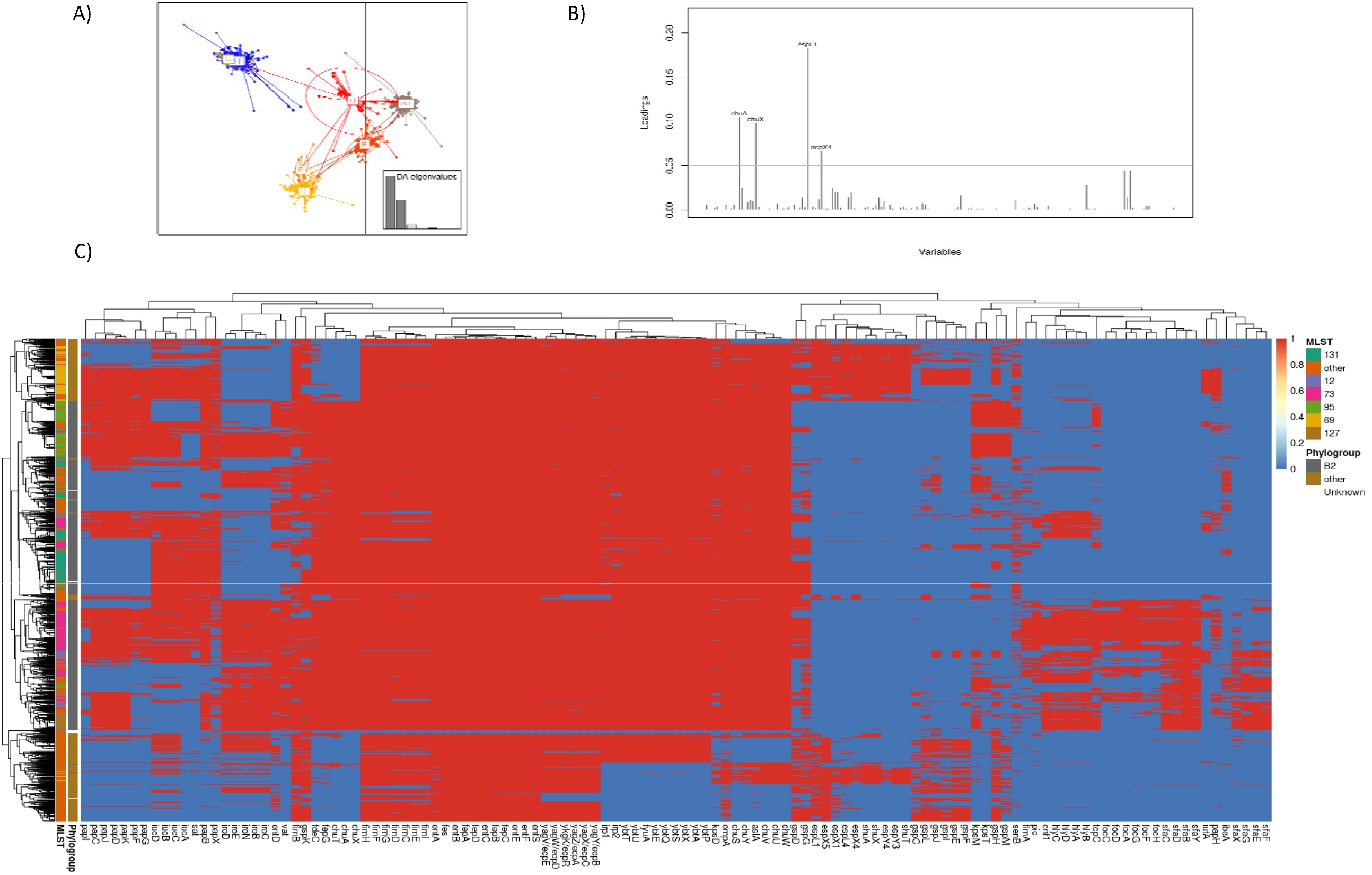
Virulence elements are structured by ST/Phylogroup in E. coli. A) Discriminant analysis of principal components plot showing clear separation of phylogroups by their virulence factor content. B) Loading plot showing genes contributing most to the discrimination of phylogroup B2 from other phylogroups. C) Heatmap of presence absence of virulence genes (x-axis) by major ST and phylogroups (y-axis).

No such structuring by virulence factors was observed for *Klebsiella* spp.; instead, virulence factors were widely distributed amongst 79 known STs and 14 isolates with no assigned ST (Fig.S8). Notably, of the 372 CA-BSIs, only 65 (17%) carried known virulence factors. There was no difference in the proportion of CA-BSI and HA-BSI isolates carrying ≥1 virulence factor (65/372 [17%] vs 92/510 [18%] respectively; p=0.9) nor carrying colibactin and/or aerobactin (15/372 [4%] vs 25/510 [5%]; p=0.7) (Fig.5).

**Figure 5:**
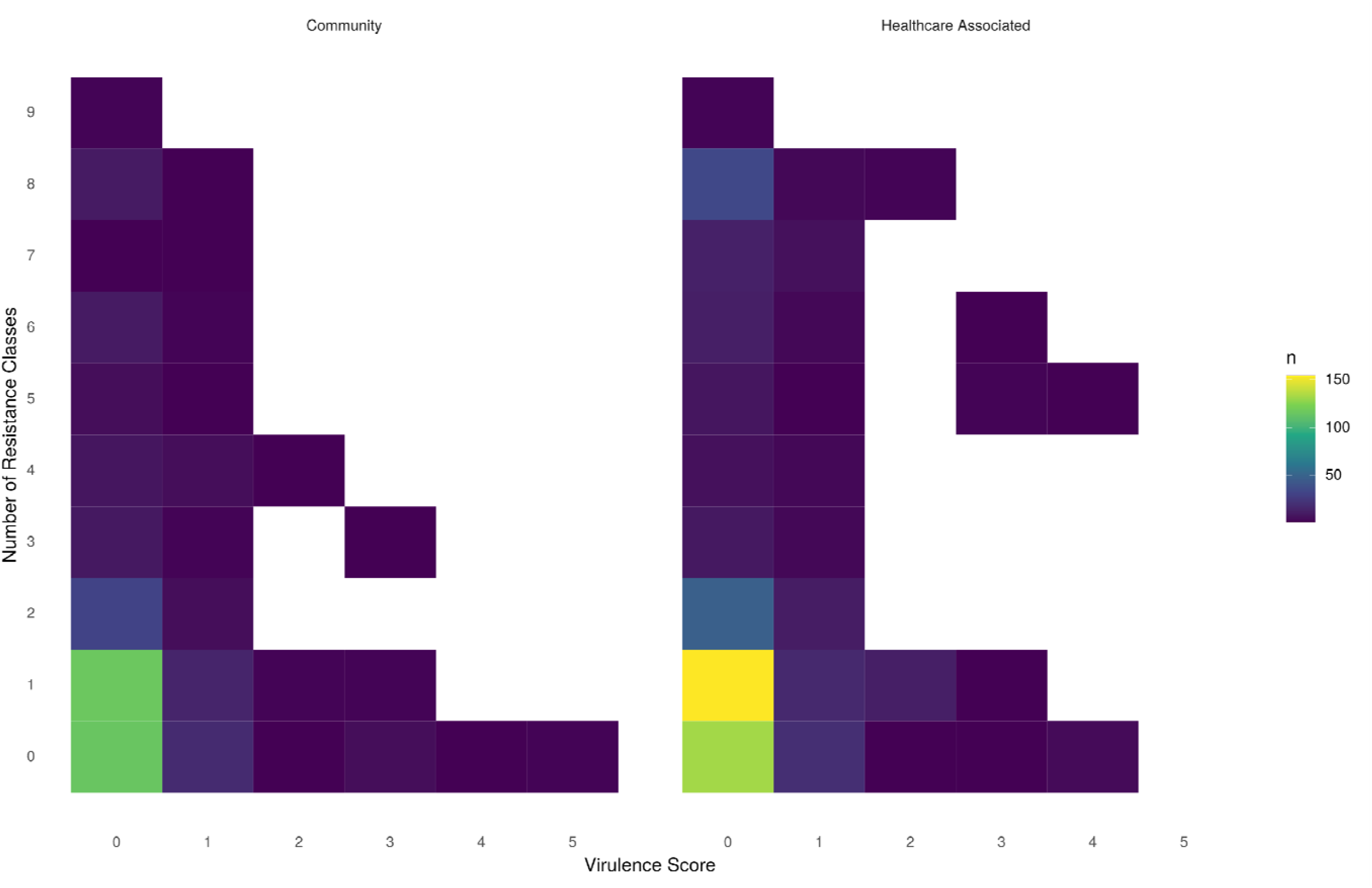
Distribution of virulence factors and antimicrobial resistance classes across community acquired (left) and healthcare associated (right) *Klebsiella* spp. isolates Virulence factor scores were assigned by Kleborate as follows: Virulence Score 0 = No acquired loci, 1= yersiniabactin, 2= yersiniabactin and colibactin (or colibactin onl = aerobactin (without yersiniabactin or colibactin), 4 = aerobactin with yersiniabactin (without colibactin) and 5 = yersiniabactin, colibactin and aerobactin.

### Plasmid replicon and plasmidome analyses reflect diversity consistent with a highly mobile accessory genome within species

Given the known importance of plasmids in the carriage and transmission of AMR genes, we sought to understand the role they might play in the relative success of major STs. Analysis of plasmid replicon profiles showed these were largely genus-restricted, with some overlap (Fig.S9). However, plasmid replicons were not structured by host lineage in *E. coli* or *Klebsiella* spp., with the exception of *K. pneumoniae* ST490 which was discriminated by the presence of the IncFIA(HI1) plasmid type (also identified in 23 *E. coli* isolates; Fig.S10).

Using our approach to predict plasmidome population structure based on k-mer similarity of contigs binned as plasmid, we observed a striking degree of plasmidome diversity within highly genomically-related isolates/STs for all species (Fig.S9); for 656 isolates with a core genome similarity >0.99 to another isolate in the dataset, the median plasmidome similarity was only 0.51 (IQR 0.28-0.86). Isolates with a near-identical plasmidome (>0.99 similarity, n=115 isolates) did however have highly similar chromosomes (median plasmidome similarity: 0.97 [IQR: 0.94-0.99]). Compared with other STs, major *E. coli* STs had larger plasmidomes (median size 106,766 vs 97,432, p<0.001) which belonged to more “plasmidome groups” (see methods) (median 3 vs 2, p<0.001). For both *E. coli* and *Klebsiella* spp., there was no evidence of different plasmid populations between those associated with HA-vs CA-BSI (p=0.4).

Finally we analysed the co-occurrence of AMR genes, plasmid replicon types and insertion sequences (ISs) within *E. coli* and *Klebsiella* spp. The networks formed were notable for the widespread co-occurrence of AMR genes and insertion sequences but not usually specific plasmid types (Figs.S11/12). This suggests that most common AMR genes are found in a diverse range of genetic contexts (e.g. multiple plasmid types, chromosomally integrated), and that horizontal gene transfer of important AMR genes in these isolates is largely facilitated by smaller, non-plasmid MGEs such as transposons/ISs, which are difficult to reliably evaluate with short-read data^50^.

## Discussion

In this unbiased longitudinal sequencing study of all (90 day-deduplicated) *E. coli* and *Klebsiella* spp. BSIs in Oxfordshire over a decade (2008-2018), we highlight the similarities and differences in the molecular epidemiology of these species. Overall, the increasing incidence of *E. coli* and *Klebsiella* spp. BSIs in Oxfordshire is not explained by the expansion of a single ST, and much of the burden (40%) of *Klebsiella* spp. disease is caused by non-*K. pneumoniae* species. Although six lineages stably account for nearly half of all *E. coli* BSIs and a clonal outbreak was observed in *K. pneumoniae*, many BSIs in our setting are caused by diverse strains with diverse accessory genomes.

We found no genomic evidence supporting the stratification of *E. coli* isolates into HA-BSI vs CA-BSI. Strains causing healthcare-associated BSIs may not be specifically healthcare-acquired but may still be healthcare-provoked (for example by the presence of indwelling devices or relative immunosuppression). Interventions targeting only HA infections (such as that of the UK government to reduce these by 50% by 2021^14^) might have limited efficacy without considering the wider ecology of these species. Whilst the incidence of *E. coli* BSIs attributed to the urinary tract increased over the study period, there was evidence that the rate of this increase was slower compared to other sources. This possibly reflects some success of infection prevention interventions such as catheter care, but also highlights the multi-faceted approach required to reduce the overall incidence rate. More work is required to understand CA-*Klebsiella* spp. BSIs because known markers of virulence found in CA disease (e.g. aerobactin, yersiniabactin, colibactin) in other studies^48^ were not seen in most of our cases. Importantly therefore, existing markers may therefore be insensitive to detect the emergence of clinically hypervirulent, CA, multidrug-resistant strains.

For *E. coli*, our analysis is consistent with previous studies demonstrating a broadly stable population structure at the MLST level with the predominance of STs 69, 73, 95 and 131. Within this however, we demonstrated some replacement in sub-clades of major STs over the study period. This may represent an adaptive strategy within major STs that allows them to continue to maintain their niche and evolve to survive changing environmental and immunological selection pressures. Differences in the presence of haem utilisation genes was a notable factor which discriminated isolates in major STs from the rest of the population, suggesting that iron metabolism might be an important selective pressure in determining invasive potential, a finding supported by a recent GWAS of a mouse model^51^. In contrast, for *Klebsiella* spp. we demonstrated that most infections are caused by sporadic STs which individually only accounted for only a small proportion of the overall population causing disease.

In *E. coli*, the incidence of isolates carrying genes conferring resistance against commonly used antimicrobial classes increased faster than those without; in general the opposite was true for *Klebsiella* spp.. The most commonly isolated MDR-ST in *Klebsiella* spp. (ST490) significantly decreased in incidence over the study period, lending credence to the idea that the relentless expansion of MDR clades is not inevitable, although it is unclear what interventions may have helped in causing its decline, particularly given that approximately a third of cases were community onset. The increasing incidence of gentamicin/ceftriaxone resistance in predominantly CA-*E. coli* BSIs seems paradoxical given that these antibiotics are rarely used in this setting. One explanation is that they are co-located on MGEs with other AMR genes encoding resistance to antibiotics more commonly used in the community (e.g. amoxicillin, trimethoprim). An alternative hypothesis might be increased exposure to third-generation cephalosporins due to the rise of ambulatory/”hospital-at-home” medical pathways where the once-a-day dosing of ceftriaxone makes it a relatively widely prescribed antibiotic in these settings. Regardless, overall, *Klebsiella* strains causing BSI appear to be exposed to declining antibiotic selection pressures compared to *E. coli*, suggesting that they are maintained and selected for in distinct ecological niches.

In *E. coli* BSI isolates, there was no genetic signal of adaptation to either healthcare or community environments, suggesting that these are not relevant niches for selection, contrary to a previous study^49^. However, this study included small numbers of isolates (n=162), and may have been unable to fully account for population structure. Furthermore our analysis suggests that, contrary to true nosocomial pathogens^52^, the ecology of the *E. coli* plasmidome is similar for HA/CA infections. Our findings support the hypothesis that the diversity observed in any given epidemiological strata (eg. age groups/infection focus/healthcare setting) is sampled from the same common ecological pool of isolates with invasive potential rather than representing specialised adaptation to any given setting. There are of course exceptions to this, such as localised outbreaks within care facilities and hospital environments.^53^ However, our data suggests that at least in Oxfordshire, these outbreaks are do not contribute significantly to the epidemiology of *E. coli* BSIs. Importantly, iatrogenic, non-pathogen-associated factors promoting invasive infection (e.g. urinary catheterisation) should be minimised to reduce the incidence of *E. coli* BSIs.

For *Klebsiella* spp., the traditional view that HA infections are opportunistic and caused by MDR isolates and CA infections are caused by isolates carrying one or more specific virulence genes (eg yersiniabactin, colibactin, aerobactin, salmochelin) appears to be an over-simplification in our setting^12,13^. The majority of CA *Klebisella* spp. infections contained none of the known genetic markers of hypervirulence, and these genes are unlikely to be a reliable method of surveillance for emerging strains with a propensity to cause invasive CA disease, at least in our setting. Whilst ESBL and MDR isolates were significantly more common in HA isolates, about a third were community onset (even using our fairly conservative definition) which significantly challenges the prevailing dogma that these are largely opportunistic HA strains.

Our study was limited by the inability to reconstruct closed genomes/plasmids using short-read sequencing data and our plasmidome analysis should therefore be interpreted with caution. The widely used PlasmidFinder database allows some inferences to be made using short read data, however there are many untypeable plasmids that are not reflected in this database. Similarly the MLplasmids classification algorithm used is trained on a limited reference set which may lead to some erroneous contig classifications. Additionally, analysis of the entire plasmidome may be too crude to identify the nuances of similarities/differences, particularly for small, shared plasmids as these represent only a small proportion of the overall plasmidome. Whilst we used a relatively conservative definition of healthcare-associated (i.e. within 30 days of hospital admission), this may have failed to capture the longer-term impacts of earlier hospital admissions.

In summary, the contrasting epidemiology of *E. coli* and *Klebsiella spp*. BSIs suggests different reservoirs, selection pressures and modes of acquisition for these genera. Separate strategies to reduce the incidence of these infections are likely to be required, and consideration of the community as a reservoir is important. The lack of reproducibility of several findings from previous studies and the poor sensitivity of current molecular markers for hyper-virulent Klebsiella surveillance highlights the critical importance of unselected sampling frames when making epidemiological inferences.

## Data Availability

Sequencing data will be uploaded to the NCBI under project accession number PRJNA604975.

## Data Availability

Sequencing data will be uploaded to the NCBI under project accession number PRJNA604975.

## Authors Notes

### Author Contributions

ASW, NS, DC and TEAP planned and managed isolate collection and acquired funding for the study. SL and KDV performed data linkage, cleaning and analysis. SL wrote the first draft of the manuscript. KC, LB, SG, JK, TD and AV performed the laboratory work. KJ and MA facilitated access to isolates and resources. ASW, SH, NS, DC and TEAP provided supervision. All authors reviewed and approved the final draft for publication.

### Data availability

All sequencing data has been deposited in the NCBI under project accession number PRJNA604975.

## Acknowledgements

We express our thanks to colleagues in the Oxford University Hospitals NHS Foundation Trust clinical microbiology laboratory who performed routine identification of the isolates in this study and catalogued then for storage. We are also indebted to Dai Griffiths who managed the isolate collected for many years. This work uses data provided by patients and collected by the UK’s National Health Service as part of their care and support. We thank all the people of Oxfordshire who contribute to the Infections in Oxfordshire Research Database.

Research Database Team: L Butcher, H Boseley, C Crichton, DW Crook, D Eyre, O Freeman, J Gearing (community), R Harrington, K Jeffery, M Landray, A Pal, TEA Peto, TP Quan, J Robinson (community), J Sellors, B Shine, AS Walker, D Waller. Patient and Public Panel: G Blower, C Mancey, P McLoughlin, B Nichols.

## References

1. Vihta, K.-D. et al.. Trends over time in Escherichia coli bloodstream infections, urinary tract infections, and antibiotic susceptibilities in Oxfordshire, UK, 1998–2016: a study of electronic health records. Lancet Infect. Dis. 18, 1138–1149 (2018).

2. English Surveillance Programme for Antimicrobial Utilisation and Resistance (ESPAUR). https://assets.publishing.service.gov.uk/government/uploads/system/uploads/attachment_data/file/843129/English_Surveillance_Programme_for_Antimicrobial_Utilisation_and_Resistance_2019.pdf (2019).

3. Public Health England. Escherichia coli (E. coli): guidance, data and analysis. GOV.UK https://www.gov.uk/government/collections/escherichia-coli-e-coli-guidance-data-and-analysis (2010).

4. Stoesser, N. et al.. Evolutionary History of the Global Emergence of the Escherichia coli Epidemic Clone ST131. MBio 7, e02162 (2016).

5. David, S. et al.. Epidemic of carbapenem-resistant Klebsiella pneumoniae in Europe is driven by nosocomial spread. Nat Microbiol 4, 1919–1929 (2019).

6. Wyres, K. L. et al.. Distinct evolutionary dynamics of horizontal gene transfer in drug resistant and virulent clones of Klebsiella pneumoniae. PLoS Genet. 15, e1008114 (2019).

7. Goswami, C. et al.. Genetic analysis of invasive Escherichia coli in Scotland reveals determinants of healthcare-associated versus community-acquired infections. Microb Genom 4, (2018).

8. Blandy, O. et al.. Factors that impact on the burden of Escherichia coli bacteraemia: multivariable regression analysis of 2011-2015 data from West London. J. Hosp. Infect. 101, 120–128 (2019).

9. Kizny Gordon, A. et al. Genomic dynamics of species and mobile genetic elements in a prolonged blaIMP-4-associated carbapenemase outbreak in an Australian hospital. J. Antimicrob. Chemother. 75, 873–882 (2020).

10. Day, M. J. et al.. Population structure of Escherichia coli causing bacteraemia in the UK and Ireland between 2001 and 2010. J. Antimicrob. Chemother. 71, 2139–2142 (2016).

11. Kallonen, T. et al.. Systematic longitudinal survey of invasive Escherichia coli in England demonstrates a stable population structure only transiently disturbed by the emergence of ST131. Genome Res. (2017) doi:10.1101/gr.216606.116.

12. Lam, M. M. C. et al.. Tracking key virulence loci encoding aerobactin and salmochelin siderophore synthesis in Klebsiella pneumoniae. Genome Med. 10, 77 (2018).

13. Holt, K. E. et al.. Genomic analysis of diversity, population structure, virulence, and antimicrobial resistance in Klebsiella pneumoniae, an urgent threat to public health. Proc. Natl. Acad. Sci. U. S. A. 112, E3574–81 (2015).

14. Wyres, K. L. et al.. Genomic surveillance for hypervirulence and multi-drug resistance in invasive Klebsiella pneumoniae from South and Southeast Asia. Genome Med. 12, 11 (2020).

15. Long, S. W. et al.. Population Genomic Analysis of 1,777 Extended-Spectrum Beta-Lactamase-Producing Klebsiella pneumoniae Isolates, Houston, Texas: Unexpected Abundance of Clonal Group 307. MBio 8, (2017).

16. SURVEILLANCE REPORT. Surveillance of antimicrobial resistance in Europe 2018.

17. Ellington, M. J. et al.. Contrasting patterns of longitudinal population dynamics and antimicrobial resistance mechanisms in two priority bacterial pathogens over 7 years in a single center. Genome Biol. 20, 184 (2019).

18. De Maio, N. et al.. Comparison of long-read sequencing technologies in the hybrid assembly of complex bacterial genomes. Microb Genom 5, (2019).

19. Hastak, P. et al.. Genomic profiling of Escherichia coli isolates from bacteraemia patients: a 3-year cohort study of isolates collected at a Sydney teaching hospital. Microb Genom 6, (2020).

20. van Hout, D. et al.. Extended-spectrum beta-lactamase (ESBL)-producing and non-ESBL-producing Escherichia coli isolates causing bacteremia in the Netherlands (2014 - 2016) differ in clonal distribution, antimicrobial resistance gene and virulence gene content. PLoS One 15, e0227604 (2020).

21. Guidance on the definition of healthcare associated Gram-negative bloodstream infections. https://www.england.nhs.uk/wp-content/uploads/2020/08/HCA_BSI_definitions_guidance.pdf.

22. Seemann, T. Shovill. https://github.com/tseemann/shovill.

23. Seemann, T. Prokka: rapid prokaryotic genome annotation. Bioinformatics 30, 2068–2069 (2014).

24. Seemann, T. abricate. (Github).

25. Lam, M. M. C. et al.. Genetic diversity, mobilisation and spread of the yersiniabactin- encoding mobile element ICEKp in Klebsiella pneumoniae populations. Microb Genom 4, (2018).

26. Wyres, K. L. et al.. Identification of Klebsiella capsule synthesis loci from whole genome data. Microb Genom 2, e000102 (2016).

27. Wick, R. R., Heinz, E., Holt, K. E. & Wyres, K. L. Kaptive Web: User-Friendly Capsule and Lipopolysaccharide Serotype Prediction for Klebsiella Genomes. J. Clin. Microbiol. 56, (2018).

28. Seemann, T. mlst. (Github).

29. Holt, K. Kleborate. https://github.com/katholt/Kleborate.

30. Beghain, J., Bridier-Nahmias, A., Le Nagard, H., Denamur, E. & Clermont, O. ClermonTyping: an easy-to-use and accurate in silico method for Escherichia genus strain phylotyping. Microb Genom 4, (2018).

31. Tonkin-Hill, G., Lees, J. A., Bentley, S. D., Frost, S. D. W. & Corander, J. Fast hierarchical Bayesian analysis of population structure. Nucleic Acids Res. 47, 5539–5549 (2019).

32. Tonkin-Hill, G. et al.. Producing Polished Prokaryotic Pangenomes with the Panaroo Pipeline. bioRxiv 2020.01.28.922989 (2020) doi:10.1101/2020.01.28.922989.

33. Seemann, T. snippy. (Github, 2015).

34. Croucher, N. J. et al.. Rapid phylogenetic analysis of large samples of recombinant bacterial whole genome sequences using Gubbins. Nucleic Acids Res. 43, e15 (2015).

35. Didelot, X., Croucher, N. J., Bentley, S. D., Harris, S. R. & Wilson, D. J. Bayesian inference of ancestral dates on bacterial phylogenetic trees. Nucleic Acids Res. 46, e134 (2018).

36. Plummer, M., Best, N., Cowles, K. & Vines, K. CODA: Convergence Diagnosis and Output Analysis for MCMC. R News vol. 6 7–11 (2006).

37. Kembel, S. W. et al.. Picante: R tools for integrating phylogenies and ecology. Bioinformatics 26, 1463–1464 (2010).

38. Arredondo-Alonso, S. et al.. mlplasmids: a user-friendly tool to predict plasmid- and chromosome-derived sequences for single species. Microb Genom 4, (2018).

39. Baker, D. N. & Langmead, B. Dashing: fast and accurate genomic distances with HyperLogLog. Genome Biol. 20, 265 (2019).

40. Kalinka, A. T. & Tomancak, P. linkcomm: an R package for the generation, visualization, and analysis of link communities in networks of arbitrary size and type. Bioinformatics 27, 2011–2012 (2011).

41. Oksanen, J. et al.. vegan: Community Ecology Package. (2019).

42. Lees, J. A., Galardini, M., Bentley, S. D., Weiser, J. N. & Corander, J. pyseer: a comprehensive tool for microbial pangenome-wide association studies. Bioinformatics 34, 4310–4312 (2018).

43. Jombart, T. & Ahmed, I. adegenet 1.3-1: new tools for the analysis of genome-wide SNP data. Bioinformatics 27, 3070–3071 (2011).

44. Csardi, G. & Nepusz, T. The igraph software package for complex network research. InterJournal vol. Complex Systems 1695 (2006).

45. StataCorp, L. P. Stata statistical software: release 16. College Station, TX: StataCorp LP; 2019.

46. Lunn, M. & McNeil, D. Applying Cox regression to competing risks. Biometrics 51, 524–532 (1995).

47. R Core Team. R: A Language and Environment for Statistical Computing. (2019).

48. Gasparini, A. comorbidity: An R package for computing comorbidity scores. Journal of Open Source Software 3, 648 (2018).

49. Eyre, D. W. et al.. Two Distinct Patterns of Clostridium difficile Diversity Across Europe Indicating Contrasting Routes of Spread. Clin. Infect. Dis. 67, 1035–1044 (2018).

50. Wick, R. R., Judd, L. M., Gorrie, C. L. & Holt, K. E. Completing bacterial genome assemblies with multiplex MinION sequencing. Microb Genom 3, e000132 (2017).

51. Galardini, M. et al.. Major role of iron uptake systems in the intrinsic extra-intestinal virulence of the genus Escherichia revealed by a genome-wide association study. PLoS Genet. 16, e1009065 (2020).

52. Arredondo-Alonso, S. et al.. Plasmids Shaped the Recent Emergence of the Major Nosocomial Pathogen Enterococcus faecium. MBio 11, (2020).

53. Brodrick, H. J. et al.. Longitudinal genomic surveillance of multidrug-resistant Escherichia coli carriage in a long-term care facility in the United Kingdom. Genome Med. 9, 70 (2017).

