## Supplement for "Molecular epidemiology of *Escherichia coli* and *Klebsiella* species bloodstream infections in Oxfordshire (UK) 2008-2018"

Supplementary Appendix

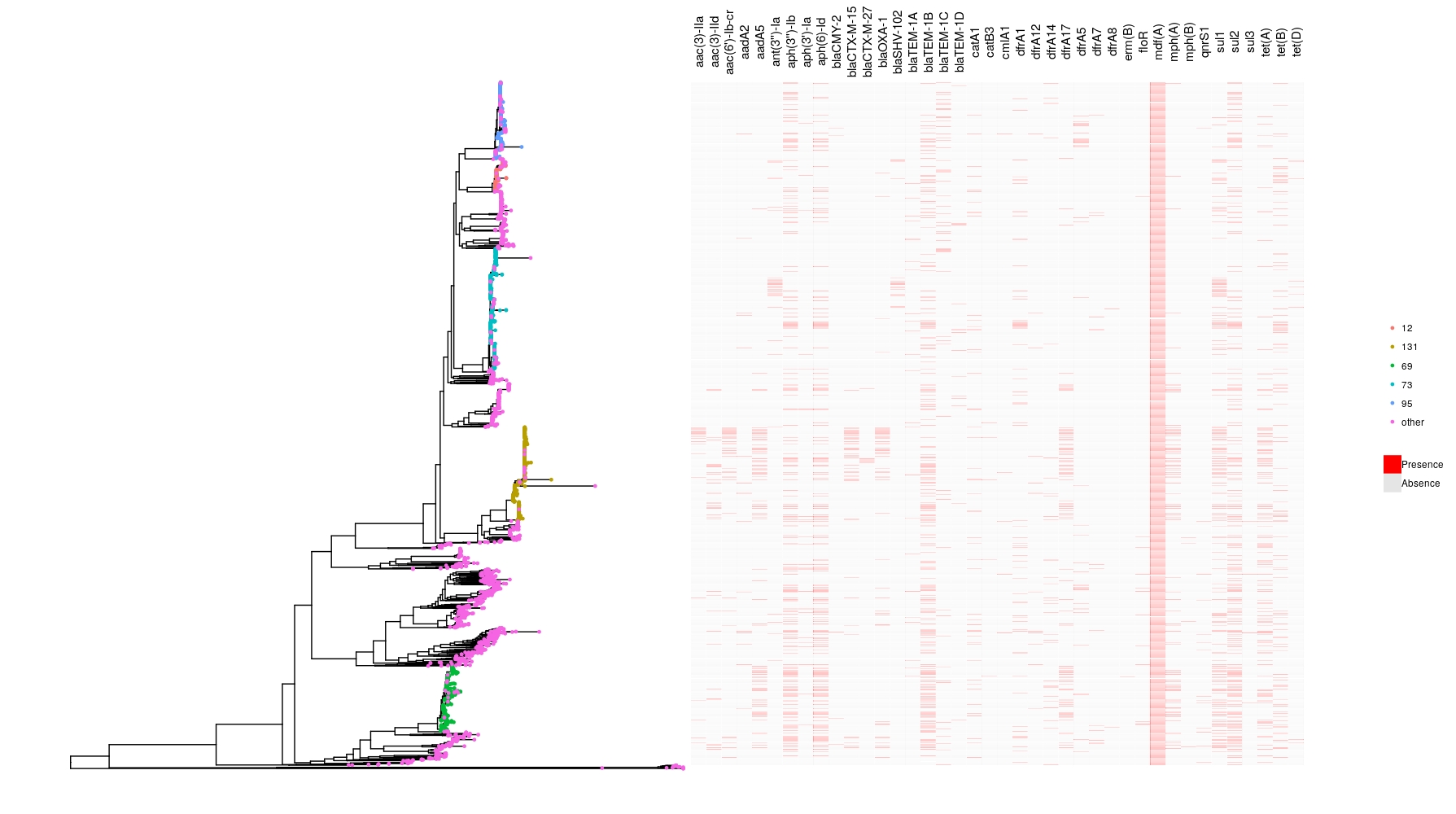

Figure S1 Gene presence/absence heatmap showing AMR gene presence/absence against the core genome phylogeny for *E. coli*. Tip colours show the location of the more common STs in the phylogeny. Only genes occurring >10 times in the dataset are shown.

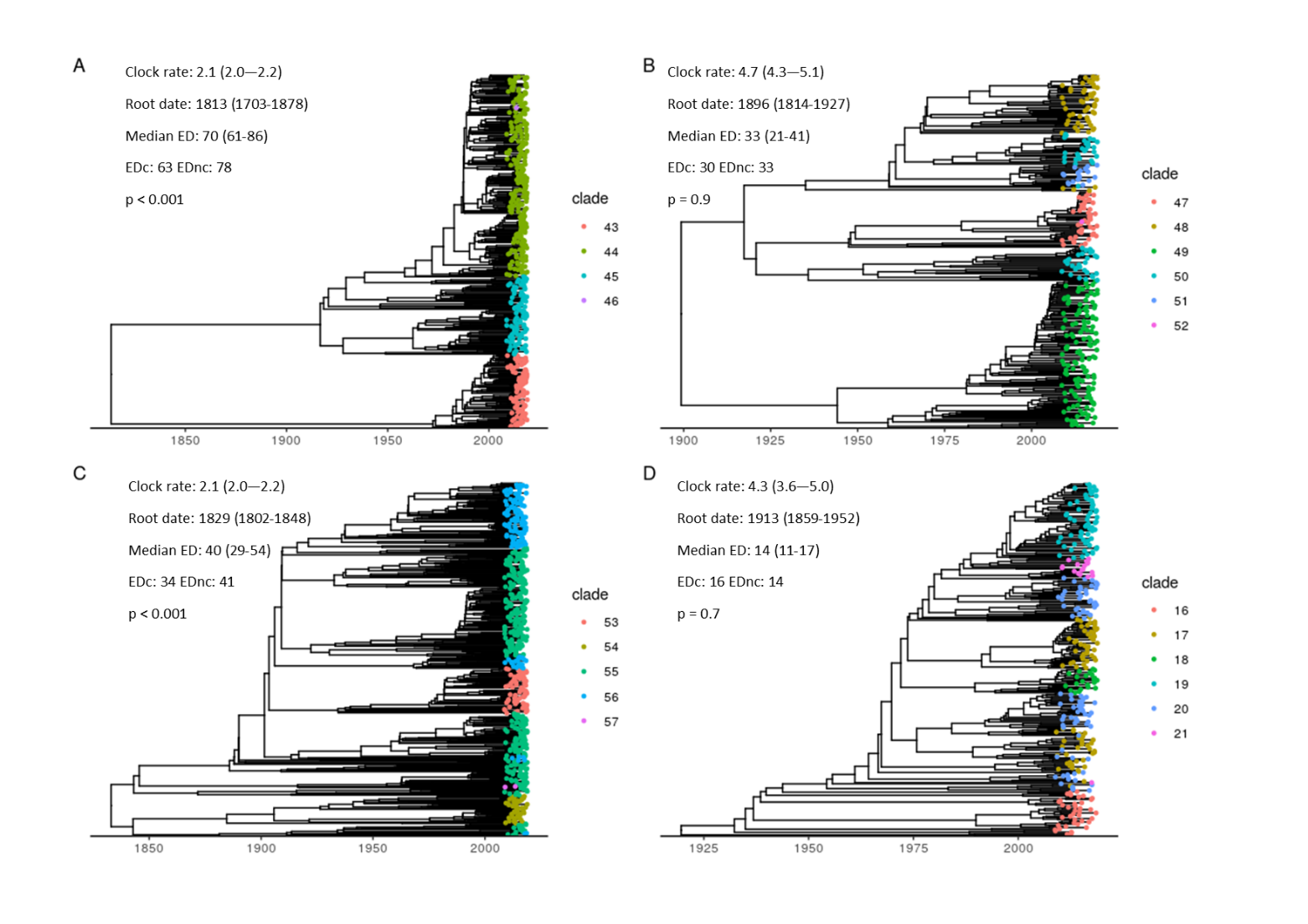

Figure S2: Time-scaled phylogenies for A) ST131 B) ST95 C) 73 and D) ST69. Tip colours represent fastBAPS clusters. The text inset shows estimates for the molecular clock (mean, 95% highest posterior density interval), SNPs/site/year), root date (mean, 95% highest posterior density interval), evolutionary distinctiveness (ED, median and interquartile range) and median evolutionary distinctiveness for isolates with (EDc) and without (EDnc) a gene conferring ceftriaxone resistance. The p value represents a Kruskal-Wallis test of the ED scores between the EDc and EDnc groups.

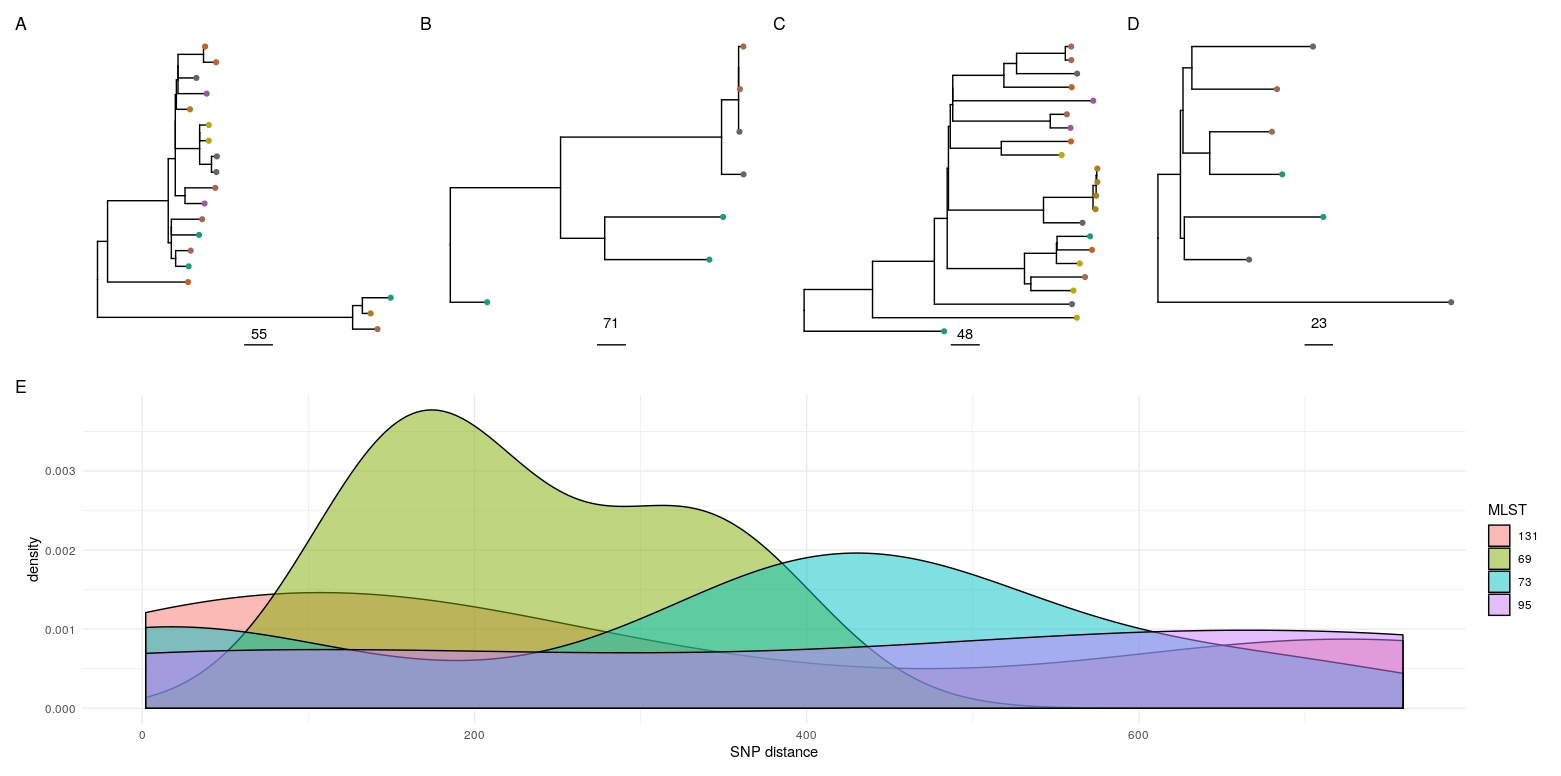

Figure S3 **–** Possible transmission within nursing homes: A-D - Phylogenies for the major STs containing isolates orginating from nursing home residents where there was more than one patient from the same GP practice in the study (the assumption being that they may therefore reside in the same nursing home). A - ST131, B - ST95, C - ST73, D - ST69. Tip colours represent GP practices. E - density plot for cophenetic SNP distances for the major ST plots shown in A-D for patients presumed to be in the same nursing home. Out of 54 total pairwise comparisons of isolates in the same ST and presumed same nursing home, 4 were <= 10 SNPs, 5 11-20 SNPs, 2 21-50 SNPs, 3 51-100 SNPs and 40 >100 SNPs.

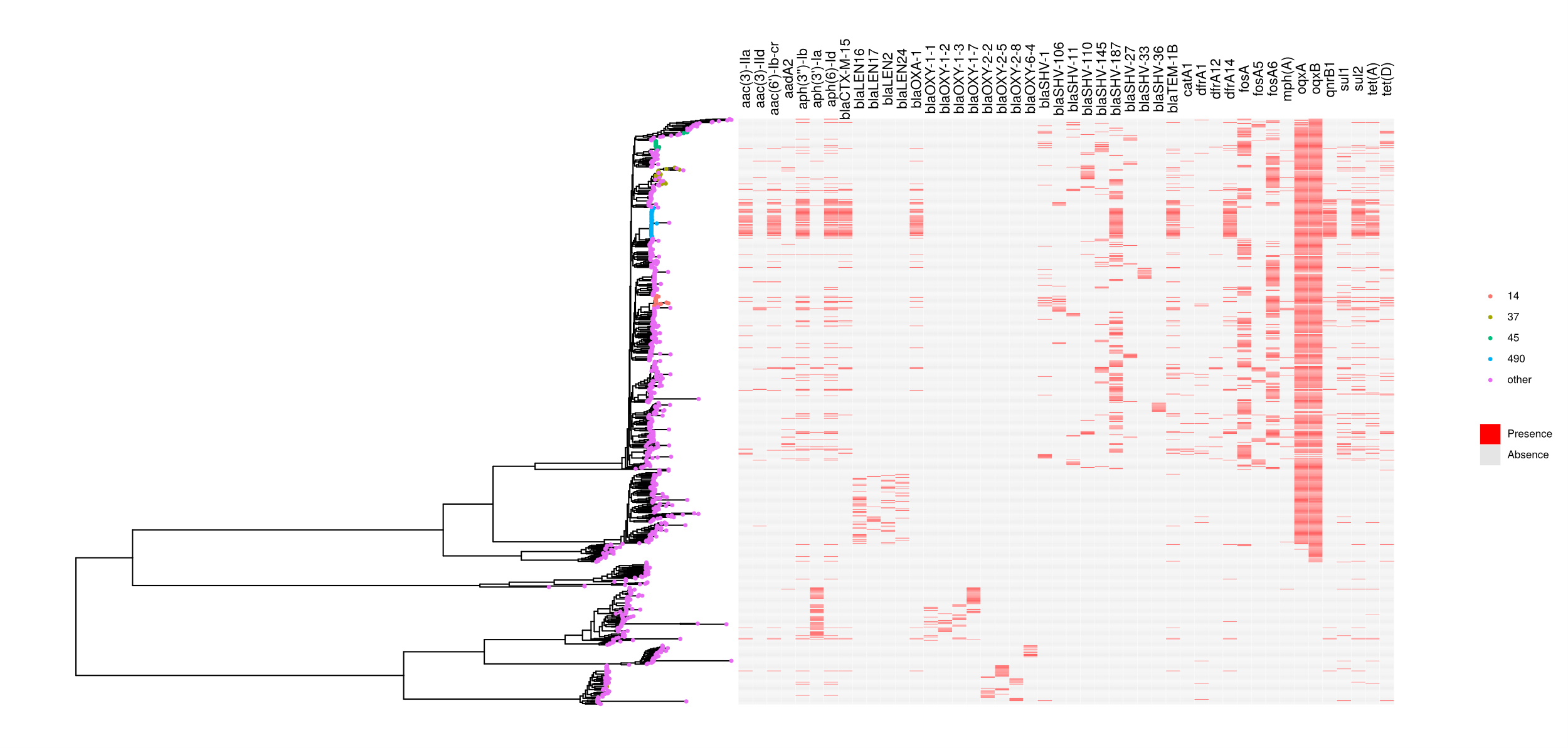

Figure S4. Gene presence/absence heatmap showing AMR gene presence/absence against the core genome phylogeny for *Klebsiella* spp. Only genes occurring >10 times in the dataset are shown. Tip colours show the location of the more common STs in the phylogeny.

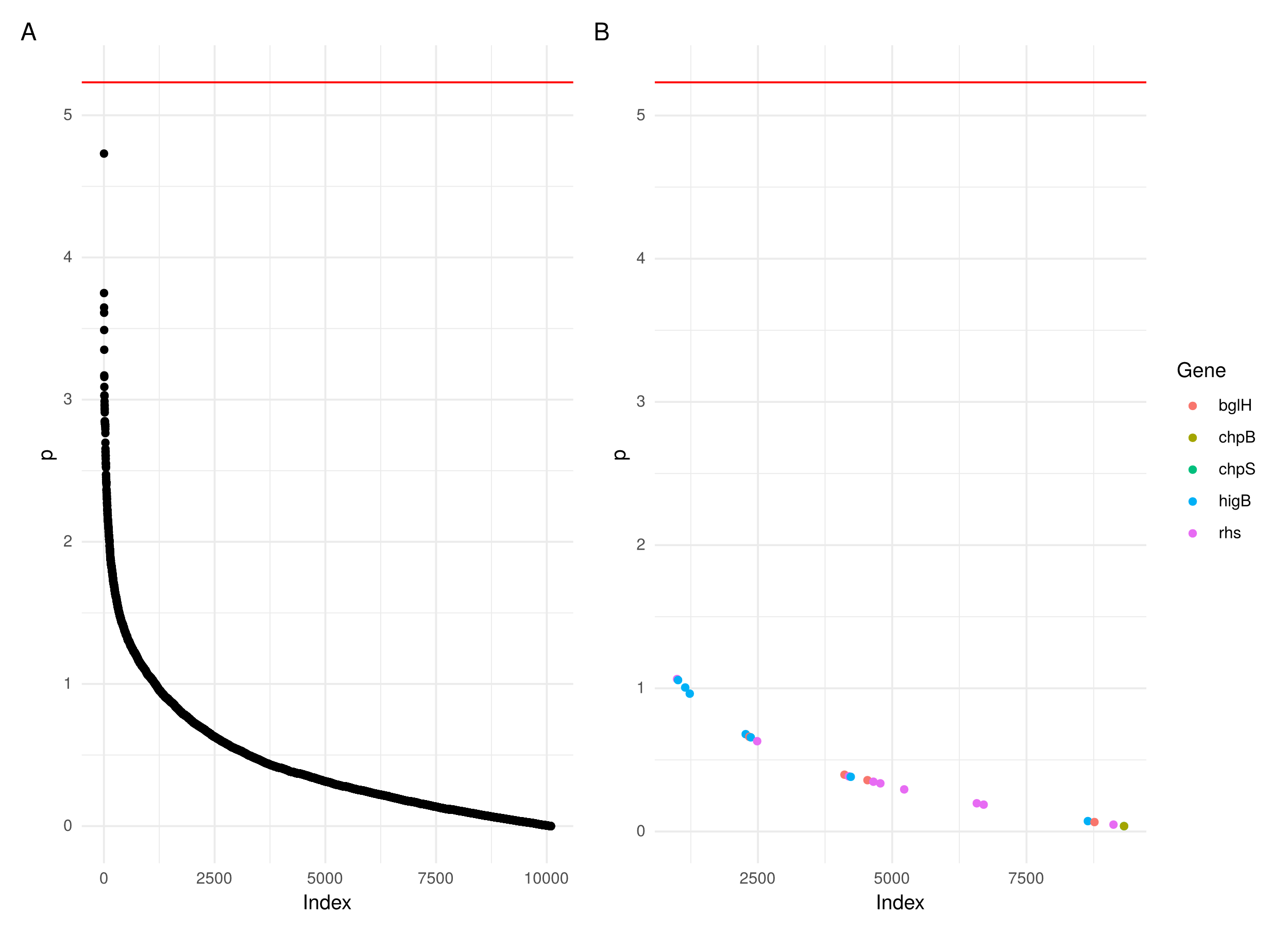

Figure S5: Manhattan plots of a pangenome wide association study of the association of genes with community/healthcare associated onset. A) all genes shown in descending order of -log10(p value). B) genes previously shown to be signficantly associated with either healthcare/community onset cases^1^. Red horizontal bar represents the Bonferroni adjusted significance threshold.

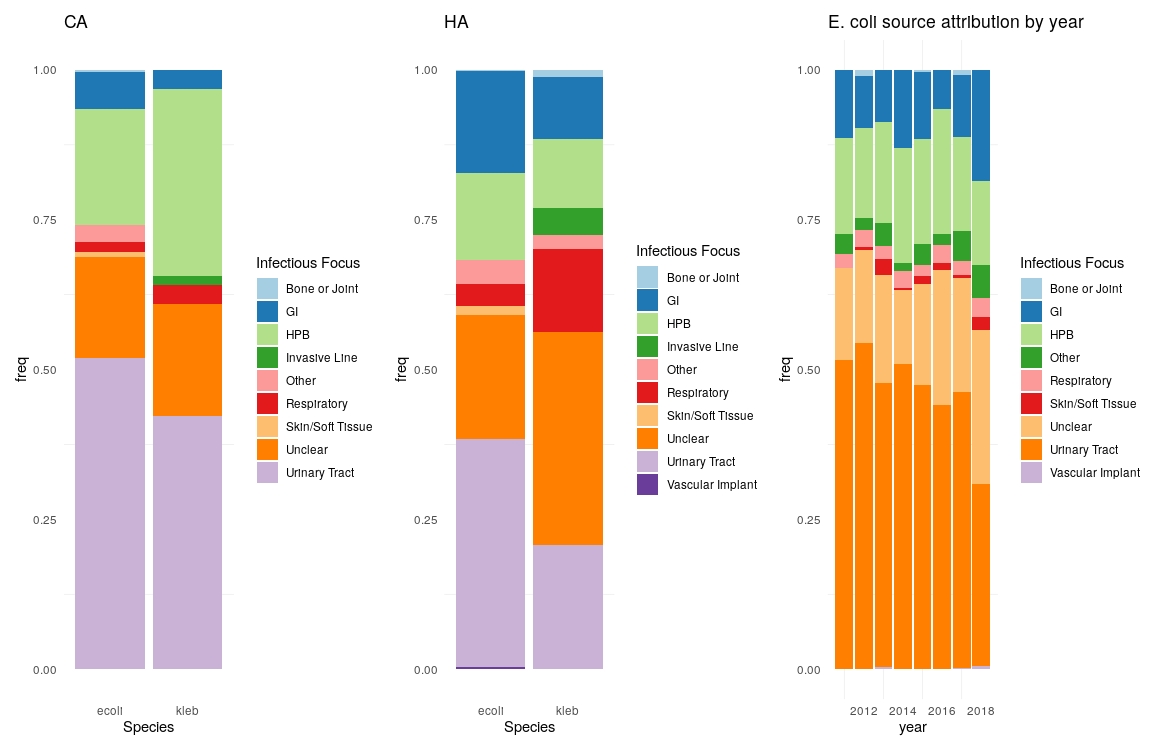

Figure S6**:** Proportions of infectious foci for CA (left panel) and HA (central panel) BSI. Source attribution by year for *E. coli* (right panel).

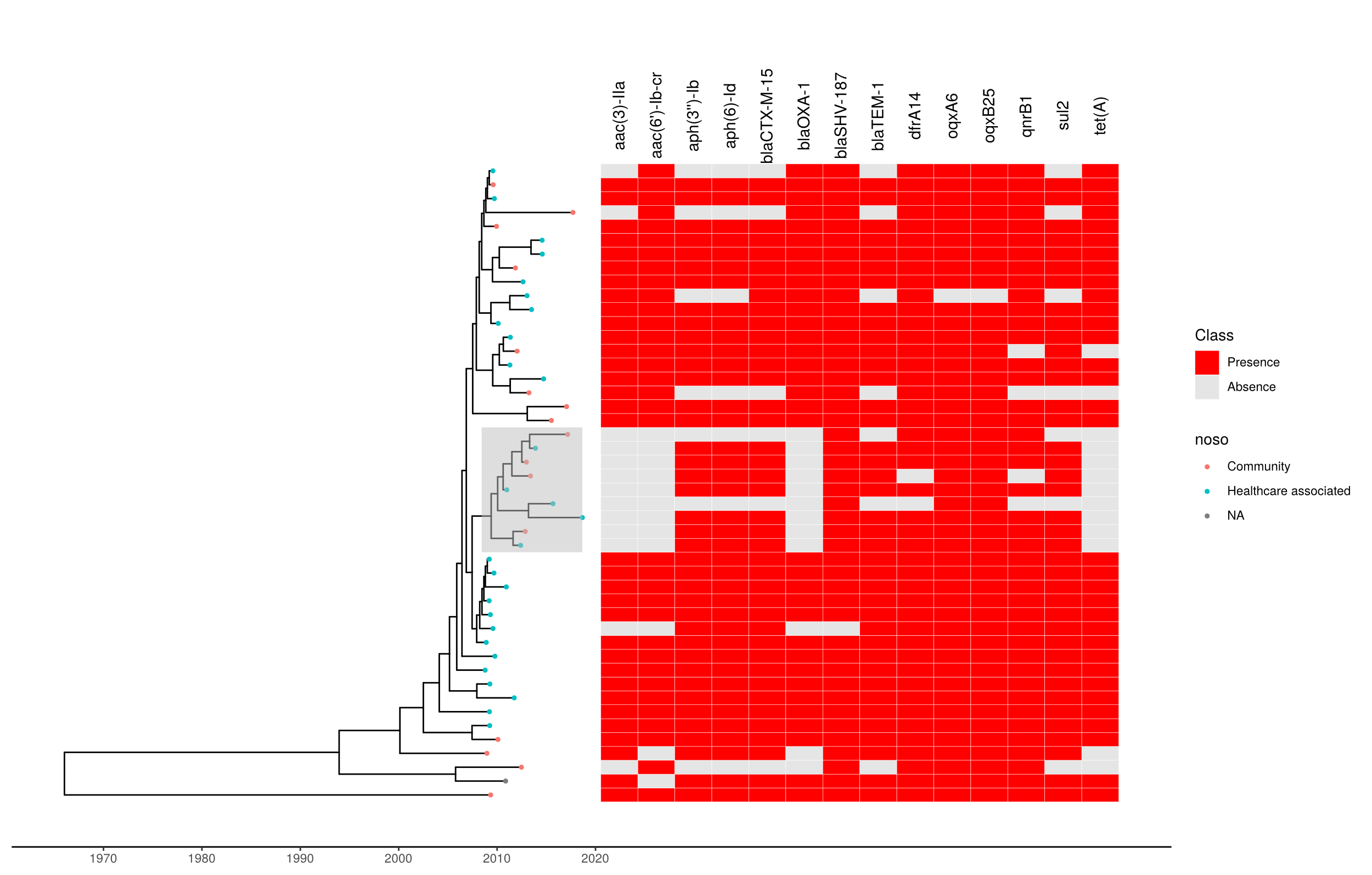

Figure S7: Timescaled phylogeny of *Klebsiella pneumoniae* ST490 with a heatmap of AMR genes. Tree tip colours indicate whether the isolate is community/healthcare associated. Only genes occurring >10 times are shown. The subclade which emerged without *aac(3)-IIa*, *aac(6’)-Ib-cr*, *blaOXA-1* and *tet(A)* is shown with a grey highlighted box. The x axis shows time in years.

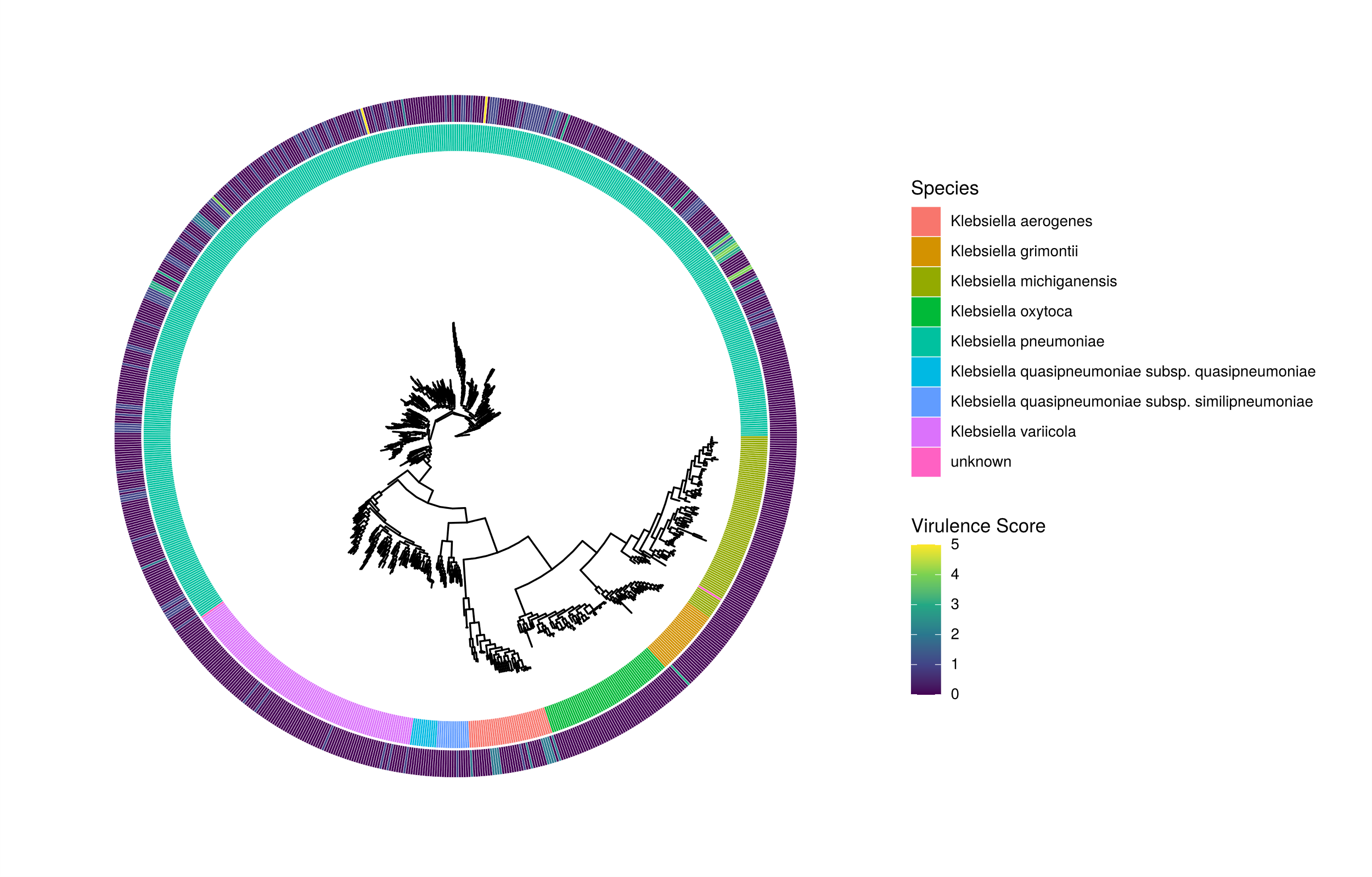

**Figure S8**: Phylogenetic tree of *Klebsiella* spp annotated with species (inner ring) and virulence score (outer ring). Virulence score is defined by Kleborate^3^ as follows: 0 – no acquired virulence loci, 1 – yersiniabactin only, 2 – yersiniabactin and colibactin (or colibactin only), 3 – aerobactin only, 4 – aerobactin and yersiniabactin (no colibactin), 5 – yersiniabactin, colibactin and aerobactin.

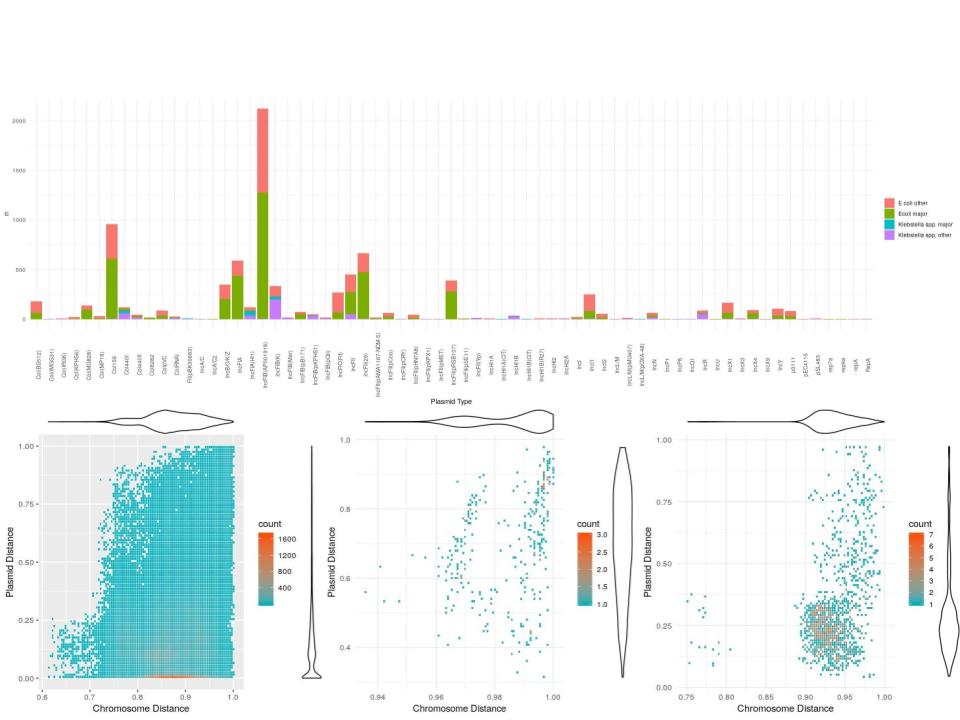

Figure S9**:** top - plasmid types in the PlasmidFinder database identified in the major/other E. coli/*Klebsiella* spp., bottom left - plot showing kmer based plasmidome similarity (y-axis) against chromosome similarity (x-axis) for isolates of the same MLST. Violin plots show the distribution of values for each axis. Origin of contigs was predicted as being plasmid/chromosome associated by ML Plasmids and distances calculated by Dashing. The bottom middle and right plots show the same thing for isolates of *K. pneumoniae* ST490 (middle) and *E. coli* ST131 (right) carrying a *bla*_CTX-M-15_ gene.

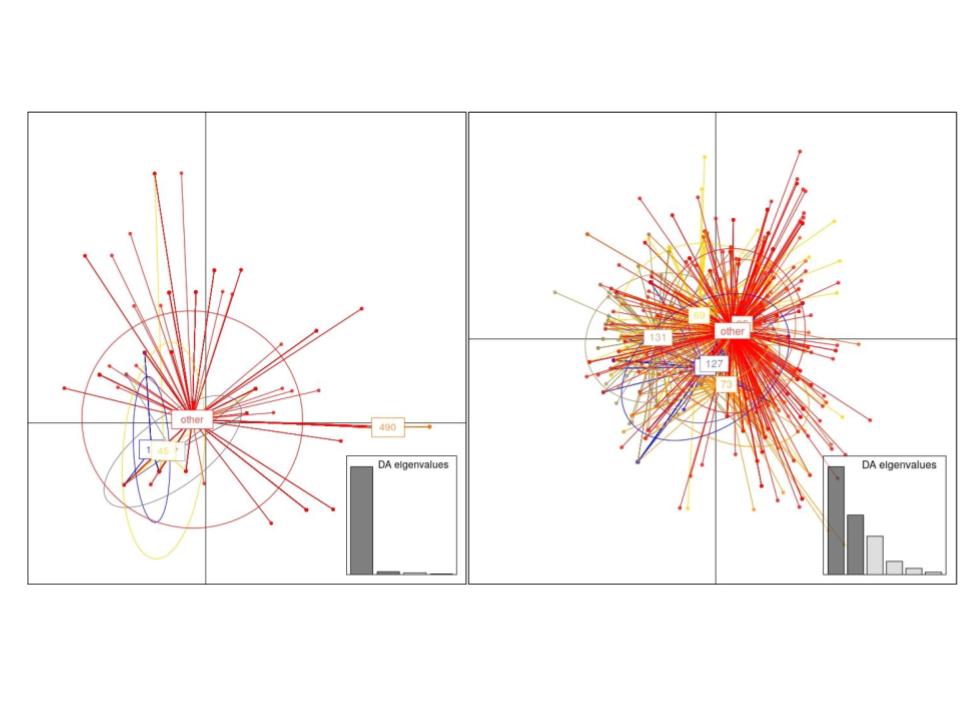

Figure S10: DAPC plots for *Klebsiella* spp. (left) and E. coli (right) showing discriminant factors which best discriminate isolates into STs based on plasmid types in the PlasmidFinder database.

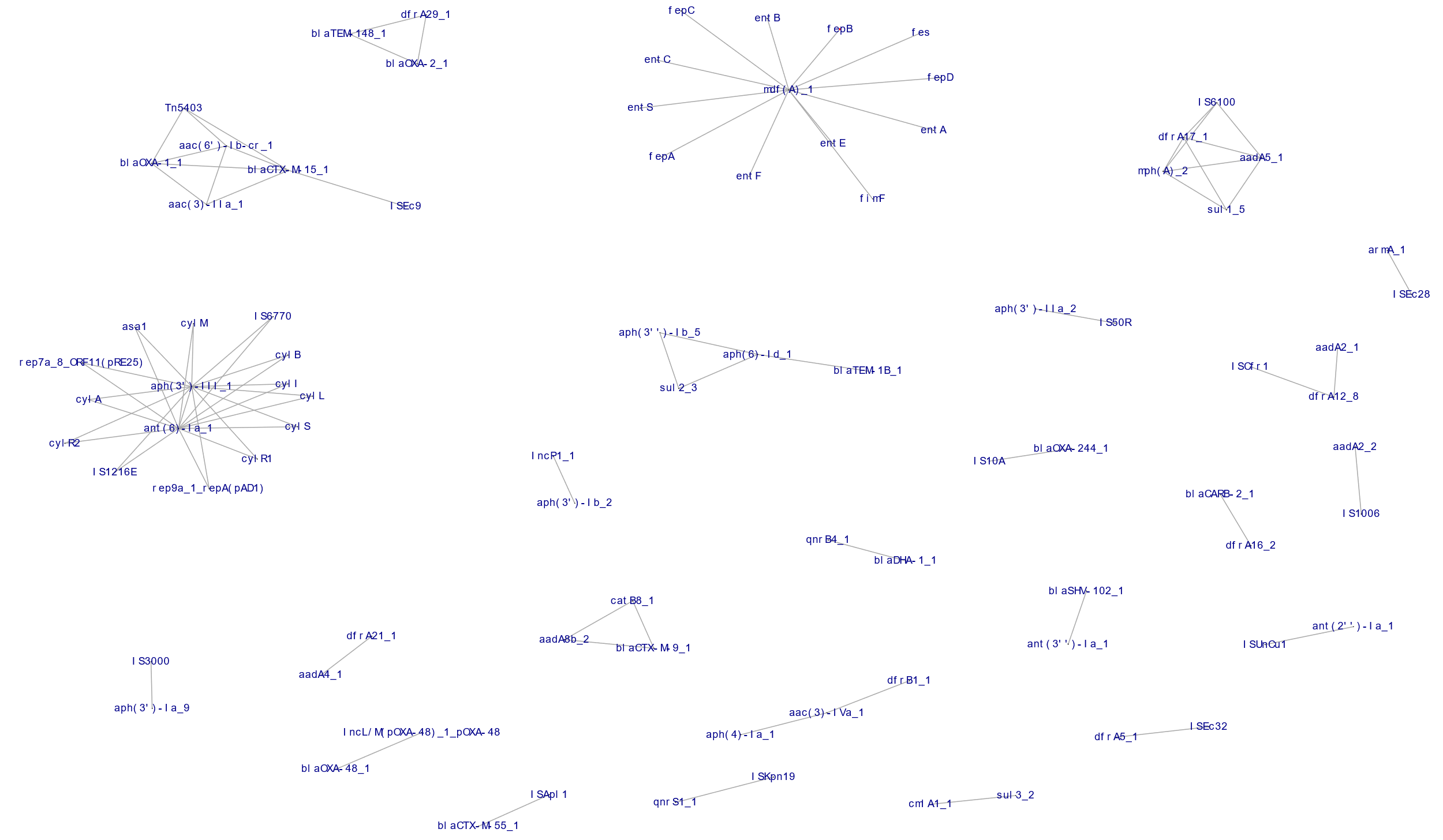

Figure S11: Networks of genes/plasmids/insertion sequences commonly co-occurring in *E. coli*. Edge lists were created from elements with a Pearson correlation coefficient >0.5.

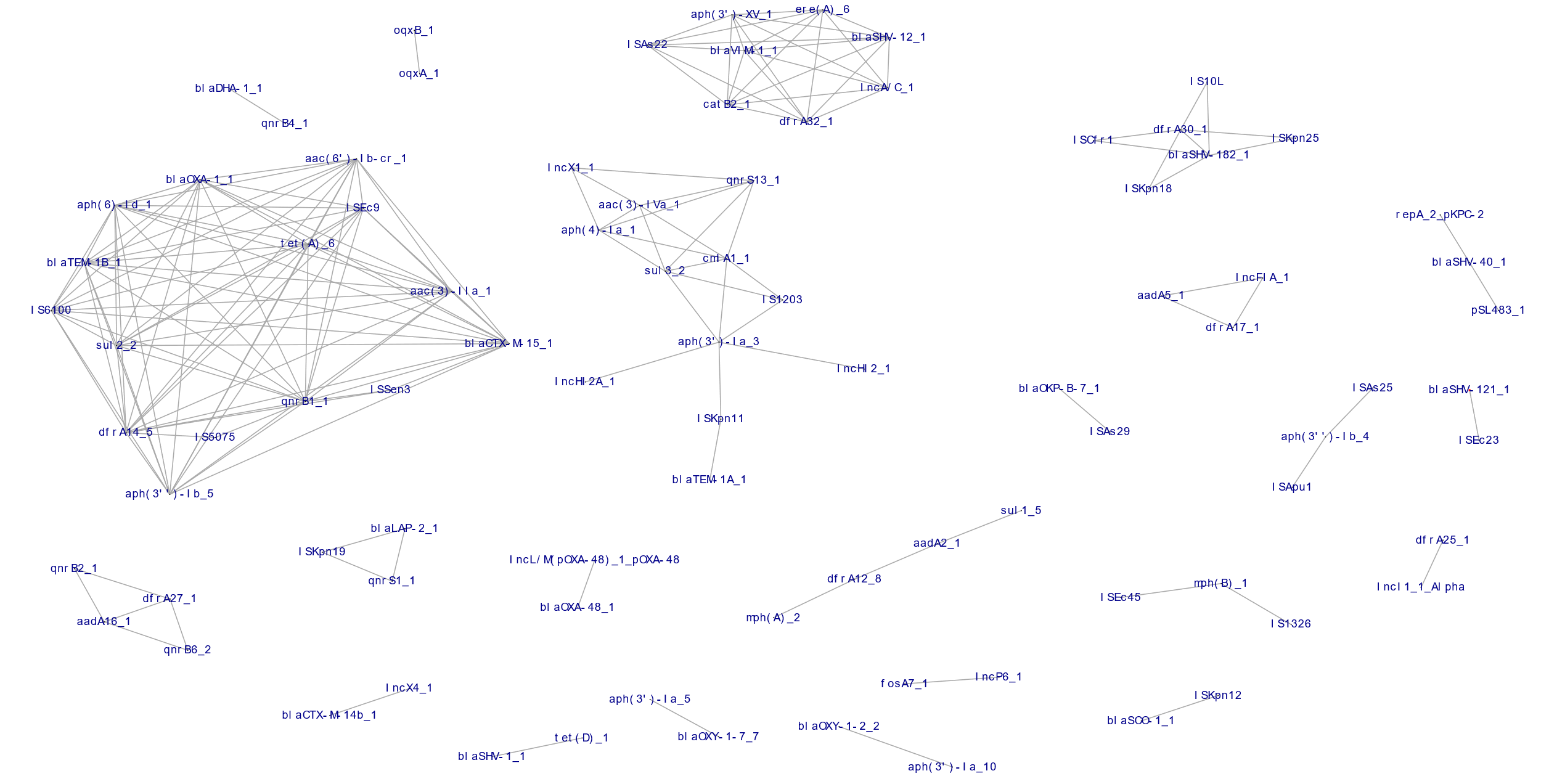
Figure S12: Networks of genes commonly co-occuring in *Klebsiella* spp. Edge lists were created from elements with a Pearson correlation coefficient >0.5.

| MLST | FastBAPS clade | IRRy | 95% CI | p_heterogeneity_ | Bonferroni corrected |
| --- | --- | --- | --- | --- | --- |
| 131 | 43 | 1.18 | 1.09-1.27 | 0.96 | 1 |
| 131 | 44 | 1.20 | 1.10-1.32 | 0.08 | 0.25 |
| 131 | 45 | 1.12 | 1.01-1.25 | 0.03 | 0.1 |
| 95 | 47 | 1.21 | 1.00-1.46 | 0.19 | 0.93 |
| 95 | 48 | 1.18 | 1.11-1.25 | 0.01 | 0.05 |
| 95 | 49 | 1.05 | 1.00-1.10 | 0.01 | 0.03 |
| 95 | 50 | 1.06 | 0.96-1.16 | 0.32 | 1 |
| 95 | 51 | 1.00 | 0.89-1.10 | 0.07 | 0.35 |
| 73 | 53 | 1.17 | 1.10-1.26 | 0.13 | 0.52 |
| 73 | 54 | 1.02 | 0.92-1.14 | 0.13 | 0.52 |
| 73 | 55 | 1.11 | 1.04-1.18 | 0.47 | 1 |
| 73 | 56 | 1.07 | 0.98-1.16 | 0.29 | 1 |
| 69 | 16 | 0.95 | 0.87-1.05 | <0.001 | <0.001 |
| 69 | 17 | 1.18 | 1.11-1.24 | 0.14 | 0.86 |
| 69 | 18 | 1.29 | 1.16-1.44 | 0.04 | 0.25 |
| 69 | 19 | 1.31 | 1.19-1.43 | 0.002 | 0.01 |
| 69 | 20 | 1.05 | 0.95-1.17 | 0.04 | 0.24 |
| 69 | 21 | 1.15 | 0.93-1.42 | 0.87 | 1 |

Table S1 – Incidence rate ratios for sub-clades of major STs identified by FastBAPS. Stacked negative binomial regression models were used to estimate incidence rate rations per year (IRRy). The p-value represents a Wald test for heterogeneity between the incidence rate for isolates of a subclade compared to the incidence rate for all other isolates in a given ST (Bonferroni coerrections to account for the number of tests performed per ST are shown to the right of this).

|  | Geographic distribution |  |  | Healthcare setting |  |  |
| --- | --- | --- | --- | --- | --- | --- |
| ST | Observed ratio | Expected ratio | p value | Observed ratio | Expected ratio | p value |
| 95 | 1.00 | 0.99 - 1.01 | 0.9 | 1 | 0.99 - 1.01 | 0.2 |
| 73 | 1.00 | 0..98 - 1.02 | 0.6 | 0.99 | 0.98 - 1.02 | 0.07 |
| 69 | 1.01 | 0.90 - 1.10 | 0.64 | 1.14 | 0.92 - 1.12 | 1 |
| 131 | 1.12 | 0.93 - 1.07 | 1 | 1 | 0.95 - 1.04 | 0.5 |

Table S2: SNP ratios (median within/between region) and (median HA/all) were calculated for each ST. Permuted distributions were created by randomising tip labels 1000 times and recalculating ratios. P values represent the number of permuted values at least as extreme as the observed value/1000. Values closer to 0 imply greater geographical/nosocomial clustering of isolates. In all cases the observed values were compatible with the null hypothesis.

| ST | Median ED CA (IQR) | Median ED HA (IQR) | p-value |
| --- | --- | --- | --- |
| 95 | 40 (31 - 49) | 41 (33 - 47) | 0.7 |
| 73 | 49 (36 - 63) | 45 ( 35 - 57) | 0.09 |
| 69 | 18 (15 - 22) | 17 (14 - 21) | 0.06 |
| 131 | 74.7 (67.4 - 91.3) | 75.5 (67.9 - 91.3) | 0.8 |

Table S3: Evolutionary distinctiveness (ED) scores for community-associated (CA) and healthcare-associated (HA) isolates amongst major *E. coli* STs. P-values represent Wilcoxon Rank Sum tests between the groups.

| Gene | Allele frequency | Likelihood ratio test p-value (adjusted for population structure) | Beta | Standard Error | Gene H^2^ | Associated source |
| --- | --- | --- | --- | --- | --- | --- |
| *papG* | 0.39 | 3.43x10^-29^ | 0.28 | 0.02 | 0.23 | Urinary tract |
| *papC* | 0.50 | 3.61x10^-20^ | 0.23 | 0.02 | 0.19 | Urinary tract |
| *papH* | 0.50 | 4.97x10^-20^ | 0.23 | 0.02 | 0.19 | Urinary tract |
| *papD* | 0.50 | 6.19x10^-20^ | 0.23 | 0.02 | 0.19 | Urinary tract |
| *papK* | 0.43 | 3.83x10^-19^ | 0.21 | 0.02 | 0.19 | Urinary tract |
| *papF* | 0.47 | 3.11x10^-16^ | 0.20 | 0.02 | 0.17 | Urinary tract |
| *papJ* | 0.40 | 2.91x10^-11^ | 0.17 | 0.03 | 0.14 | Urinary tract |
| *papE* | 0.32 | 1.86x10^-9^ | 0.15 | 0.03 | 0.13 | Urinary tract |

**Table S4**: Top hits from a pangenome-wide association study (PGWAS) performed using Pyseer^2^ of the assocation of gene presence/absence with pysician identified BSI source. Shown are the first 8 hits in order of likelihood ratio test p value. The bonferonni adjust significance threshold was 1.10x10^-6^.

| ST131 | HG941718.1 |
| --- | --- |
| ST95 | NZ_CP012625 |
| ST73 | AE014075.1 |
| ST69 | ERR3986909 |
| ST490 | accession to follow, custom de novo assembly |

Table S4: Accession numbers of reference sequences used

**References**

1.	Goswami, C. *et al.* Genetic analysis of invasive Escherichia coli in Scotland reveals determinants of healthcare-associated versus community-acquired infections. *Microb Genom* **4**, (2018).

2.	Lees, J. A., Galardini, M., Bentley, S. D., Weiser, J. N. & Corander, J. pyseer: a comprehensive tool for microbial pangenome-wide association studies. *Bioinformatics* **34**, 4310–4312 (2018).

3.	Holt, K. Kleborate. https://github.com/katholt/Kleborate.
